# Mapping of person-centred sexual and reproductive health interventions at primary healthcare settings: An umbrella review

**DOI:** 10.1101/2025.09.16.25335742

**Authors:** Ghada E. Saad, Sohayla El Fakahany, Lara Abi Jaoude, Petra Halawi, Zeina Karam, Tamar Kabakian-Khasholian, Faysal El Kak, Stephen J. McCall

**Author notes:** Senior author (Faysal El Kak). Corresponding author: Stephen J. McCall, Center for Research on Population and Health, Faculty of Health Sciences, American University of Beirut, Beirut 1107 2020, Lebanon.

## Abstract

**Introduction:** Sexual and reproductive health (SRH) is an essential component of primary healthcare (PHC). Advancing person-centred SRH promotes gender equality and ensures women and girls access high-quality care that respects their autonomy. This umbrella review synthesises key findings on existing person-centred SRH interventions in primary care settings, globally.

**Methods:** Searches were conducted in five bibliometric databases from 2016-2024. Reviews were included if they referred to person-centred SRH interventions, aimed at enhancing women’s autonomy, in PHCs. Two researchers independently screened reviews for inclusion, extracted and appraised quality of eligible reviews. Data was synthesised to identify themes and related person-centeredness.

**Results:** Of 1,300 identified reviews, 21 were included (twelve scoping, seven systematic, one integrative, and one narrative review). Geographic scope varied where eight were global, six focused on low- and middle-Income countries, and seven on high-income countries. These reviews explored multiple SRH topics (e.g., family planning, STIs/HIV, breast/cervical cancer). Four reviews explicitly referred to person-centredness; otherwise, person-centeredness was inferred. Synthesis identified five themes:1-Interactive decision- and counselling-support tools;2-Provider-initiated clinical prompts/support; 3-Remote/digital information channels;4-Integrated service delivery and accessibility strategies; and 5-Social/peer-led navigation. These interventions mainly addressed person-centred care domains of autonomy, dignity and communication, while trust, social support, and facility environment were addressed less.

**Conclusions:** Person-centred SRH interventions in PHCs have varied delivery modes highlighting innovation and a shift towards SRH care that is more responsive, accessible and respectful. Nevertheless, given limited explicit evaluation of person-centredness, future interventions need to encompass person-centred components to achieve holistic, person-centred SRH care for women and girls.

**SUMMARY BOX - Key Messages:** *What is already known about the topic:* There are numerous existing systematic and scoping reviews on Sexual and Reproductive Health (SRH) interventions tackling specific SRH services. Despite this growing body of evidence and many reviews on SRH services and interventions, the existing literature is fragmented and often fails to address person-centeredness of these services and interventions. To our knowledge, no previous umbrella review has synthesised the available evidence and specifically addressed the concept of person-centred SRH care within primary healthcare settings.

*What this study adds:* This umbrella review provides a comprehensive overview of person-centred SRH interventions by synthesising high-level findings from 21 eligible reviews. By collating a global collection of systematic and scoping reviews on various SRH topics, this study highlights the available SRH interventions, describing their person-centredness, a concept that is not widely emphasised in the existing literature. It presents a high-level narrative synthesis, offering a comprehensive overview of the available person-centred SRH interventions in primary care settings that have the potential to enhance girls’ and women’s autonomy.

*How this study might affect research, practice and policy:* This umbrella review underscores the limited emphasis on person-centred SRH interventions, and the need to design more holistic, tailored interventions that consider the multiple domains of person-centred care to ensure women receive respectful SRH services that ensure autonomy, choice and supportive care. The synthesis of results highlights the need for standardised measurements of clinical and person-centred care outcome indicators.

## INTRODUCTION

Sexual and reproductive health and rights (SRHR) are fundamental aspects of people’s health and wellbeing.^1^ Investing in comprehensive sexual and reproductive health (SRH) care is essential for achieving universal health coverage, gender equity and empowerment.^2^ In 1994, the International Conference on Population and Development emphasized the importance of individual rights, gender equality and reproductive health.^3^ Significant progress has been made in improving the quality of SRH care, nevertheless, further work is required to ensure comprehensive SRH care is present in primary healthcare settings.^4^

Person-centred care (PCC) emerged as a prominent aspect of quality, holistic healthcare, prioritising individual needs, preferences, and experiences.^5-7^ Proper implementation of PCC is essential for improving SRH care quality^5^ and strengthening health systems to respond to diverse challenges from humanitarian crises to ageing populations.^8^ While a single definition of person-centred SRH care is lacking,^7^ Sudhinarset et al (2017), defined person-centred reproductive health care as consisting of several domains, not necessarily mutually exclusive, that would lead to better care quality: dignity, autonomy, privacy & confidentiality, communication, social support, supportive care, trust and health facility environment.^5^ Though PCC and patient-centred care are often used interchangeably,^6 9^ PCC focuses on the individual’s context and preferences, holistically, rather than defining them by their disease.^9^

SRH is a key component of Primary Health Care (PHC), which serves as the first point of clinical contact and the foundation for universal health coverage by ensuring affordable, accessible and non-discriminatory high-quality services.^10^ Person-centred SRH is essential for all individuals regardless of sex and gender; however, women and girls disproportionately face adverse SRH outcomes as a result of context-specific socio-cultural or systemic barriers to care.^1 11^ SRHR frameworks heavily emphasise women’s empowerment, gender equity and self-agency as a priority.^11 4^There is a growing body of evidence supporting person-centred SRH care, and numerous reviews of SRH interventions implemented within PHC settings, generally and among particular populations such as adolescents or refugees.^9 12 13^ Nevertheless, this literature is fragmented and rarely refers to the person-centredness of interventions. To fill this gap and align with SRHR frameworks that highlight existing inequalities, this umbrella review aims to identify person-centred interventions for SRH care and education that enhance women’s autonomy and have been implemented in PHC settings, through a high-level synthesis of key findings from published reviews.

## METHODS

The umbrella review was structured in accordance with the PRIOR (Preferred Reporting Items for Overviews of Reviews) reporting guidelines.^14^

### Eligibility criteria

The inclusion and exclusion criteria are described using the PICO (Population, Intervention, Comparator, and Outcome) and PICo (Population, Intervention, Context) research question elements.^16^ Given the presence of different types of reviews, PICO criteria were used for reviews that assessed intervention outcomes, and PICo characteristics for reviews that only described interventions without comparing them to any other type of intervention or lack of intervention. The inclusion and exclusion criteria were based on the following details.

#### Population

The population of interest was adolescent girls and women, without any specific age range. Boys and men receiving person-centred SRH interventions were included when such interventions led to women’s SRH self-agency and autonomy. Reviews that did not include this population were excluded and interventions/primary studies within the reviews that did not fulfil the criteria were not extracted. When there was no disaggregation by gender, the intervention was included.

#### Intervention

Person-centred SRH interventions delivered in PHC settings were included in the umbrella review. PCC was based on Sudhinaraset et al (2017)’s definition, which consists of eight domains: dignity, autonomy, privacy & confidentiality, communication, social support, supportive care, trust and health facility environment.^5^ When not explicitly stated, person-centredness of SRH interventions were inferred from one or more of these domains. Interventions in eligible reviews that did not align with any PCC domains or PHC setting were not extracted. Further details are found in the Supplemental Material-Detailed Methods.

#### Comparison

For reviews that assessed intervention outcomes, any comparator was considered. Reviews with no comparator were also considered if they presented person-centred SRH interventions.

#### Context

To be included, reviews had to describe SRH interventions that were implemented within PHC settings. Interventions delivered in other settings, or were that social media-based were excluded. All geographic locations were included.

#### Outcomes

Included outcomes ranged from clinical outcomes such as increased uptake and continuation of care, to PCC outcomes, such as, self-agency, autonomy, increased satisfaction, service user/provider interaction and communication, mental well-being, increased knowledge. This list of outcomes is not exhaustive and was kept broad to inlcude outcomes that each eligible review defined. Outcomes related to intervention cost evaluations were excluded.

#### Additional elements

Any type of literature review was eligible for inclusion. The time frame of interest was from 2016 to present. No language restrictions were applied.

### Information Sources

On September 15, 2024, five electronic bibliographic databases were searched: MEDLINE (Ovid), CINAHL Ultimate (EBSCOhost), EMBASE (Elsevier), Scopus (Elsevier) and Cochrane Database of Systematic Reviews (Cochrane Library).

### Search Strategy

Search terms were based on four concepts: SRH topic areas, intervention terms, PHC context and reviews. Medical Subject Headings (MeSH) controlled vocabulary and keywords for each concept to ensure comprehensiveness of the search (Further details in Supplemental Material-Detailed Methods and Table S1).

### Selection Process

The search results were exported into Endnote and duplicates removed. The results were then exported into Covidence Systematic Review software, where more duplicates were removed, and the software was used to managing the screening and data extraction processes. Inclusion and exclusion screening criteria were developed for the title/abstract and full-text screening stages and entered into Covidence software. Prior to beginning the screening process, four researchers (SEF, LAJ, PH, ZK) conducted a calibration exercise to pilot the screening criteria and discussed inconsistencies until there was consensus.

Subsequently, the researchers independently screened titles/abstracts and full-text articles for inclusion, in pairs. Disagreements were resolved through discussion, and when the pair of researchers did not reach a consensus, a third researcher (GES) was consulted. Furthermore, there was no exclusion of primary studies or interventions that overlapped between the eligible reviews.

### Data collection process

A data extraction form was developed including sections on the review aims, methods, and research elements (Supplemental Material-Detailed Methods). Data was extracted from each eligible review, independently by two researchers (SEF & LAJ or PH & ZK). Any discrepancies were resolved through discussion, and a third researcher (GES) was consulted when needed. A third reviewer (GES) validated the final extracted data. To identify the degree of overlap between primary studies in eligible reviews, a citation matrix was created listing the relevant primary studies in each eligible reviews. Subsequently, the degree of overlap was measured using the corrected covered area (CCA) formula and the overlapping primary studies were assessed.^18 19^

### Data Items

All clinical and person-centred outcomes (defined above) that were presented in the synthesised results of eligible reviews were included. The effect measures were reported as presented eligible reviews. The reported outcome measurements included odds ratios, prevalence ratios, proportions, or narrative presentations.

### Risk of Bias Assessment

The quality of eligible reviews was assessed using the AMSTAR 2 critical appraisal tool for systematic reviews, which includes randomized or non-randomized studies of healthcare interventions.^20^ The researchers (SEF, LAF, PH, ZK) independently assessed the eligible reviews, in pairs. The overall rating was assessed based on satisfying the checklist’s critical and non-critical criteria, as described by Shea et al (2017).^20^ The quality of primary studies was assessed based on information collected from the eligible reviews on the risk of bias. Lack of quality assessments were documented.

### Synthesis Methods

Extracted data was synthesised narratively. This was planned given the high heterogeneity in interventions and outcome measures in the eligible reviews. Moreover, since all types of reviews were included, meta-analysis was not possible. In the narrative synthesis, reviews and interventions were grouped based on the topics and type of interventions. The overall positive and negative outcomes for each group of interventions were explored and the person-centred domains that best addressed the interventions were identified by the researchers.

### Reporting Bias Assessment and Certainty Assessment

To document risk of bias due to missing results arising from reporting biases, one researcher (GES), collected related information from the eligible reviews, and subsequently, summarized the findings and potential gaps. Certainty assessment for outcomes of interest was also collected by the researcher (GES) from the eligible reviews when present. For both assessments, there was no re-assessment conducted given the primary aim of the umbrella review was to identify existing interventions rather than assess effectiveness.

### Patient and Public Involvement

Key stakeholders were involved in the design of the research questions and methodology of the umbrella review.

### Ethics Statement

This work did not include human participants and did not require ethical approval from the Institutional Review Board at the American University of Beirut.

## RESULTS

The search identified 2,347 records. After duplicates were removed, 1,300 records were screened resulting in 21 reviews being included (See Figure 1). These included seven systematic reviews, twelve scoping reviews, one narrative review and one integrative review. The most common reasons for full-text exclusion were: not explicitly an SRH intervention or not a type of review.

### Characteristics of eligible reviews

The eligible reviews covered several areas of SRH, mainly, family planning, STIs and HIV, breast & cervical cancer and sexual health (Figure 2). Target populations were generally broad: ten reviews did not focus on any specific population, two reviews focused on adolescents, one review looked at all women primarily and adolescents secondarily, one review focused on reproductive aged population receiving care from clinic settings, four reviews focused on reproductive aged women, two reviews were aimed at urban populations, one review studied women residing in rural settings, one review focused on migrants and one review targeted conflict-affected populations. The reviews were published between the years 2017 and 2024 (Figure S1) with the highest frequency in 2021 (five reviews). The geographic scope of the reviews was somewhat varied. Eight reviews had a global scope, six focused on low- and middle-income countries and seven were in high-income countries. The geographic distribution of primary studies is illustrated in Figure 3, showing that most primary studies took place in North America. Table S2 presents a description of the characteristics of each eligible review.

### Primary study overlap

Primary study overlap was minimal and the CCA recorded 0.12% (seven overlapping studies across 306 total studies). Four primary studies overlapped in two of the reviews ^25 27^ and the content was specific to the description of three interventions (MyPath, OKQ, and FPQ/RepLQ). Three other duplicates were identified: two reviews^25 26^ had one common primary study about the ‘My Birth’ intervention, two reviews^27 31^ had a common study on the Reproductive Health Life Plan (RFLP) intervention, and two reviews ^34 35^ discussed the same intervention on patient navigation support. The overlap was documented and did not impact the synthesis.

### Risk of bias in eligible reviews and primary studies

Confidence in the quality of eligible reviews, based on the AMSTAR 2 quality appraisal checklist^20^ was rated as moderate for one review^21^, low for three reviews^12 30 31^ and critically low for 17 reviews.^22-29 32-40^ Of the critical domains, 7/21 reviews had protocols, most adhered to a partial comprehensive search strategy (mainly did not search reference list of included studies) and only one review provided a list of excluded articles with justifications. Only 8/21 assessed risk of bias (mainly using mixed methods study assessment tools) and five of these reviews used these assessments to interpret results. No meta-analyses were conducted. Further information on AMSTAR2 critical and non-critical domains by review are presented in Table S3.

### Synthesis of results

The findings were synthesised based on the mode of delivery and purpose of the intervention into five themes: 1-Interactive decision- and counselling-support tools; 2-Provider-initiated clinical prompts and support; 3-Remote and digital information channels; 4-Integrated service delivery and accessibility strategies; and 5-Social and peer-led navigation. Further details about interventions extracted from eligible reviews are described in Table S4.

#### 1. Interactive decision- and counselling-support tools

A significant body of evidence described interventions designed to facilitate shared decision-making, as shown in Table 1-Theme 1. These interventions included digital tools shared with service users while in the waiting room to assist in identifying family planning preferences, needs, and concerns prior to the counselling session, coaching support by an educator in the waiting room, and counselling techniques that aimed to enhance the user-provider dynamic towards a collaborative relationship. These interventions primarily touched on the PCC domains of autonomy, dignity, communication, and supportive care, since they are specifically designed to facilitate shared decision-making and tailor sessions to user preferences. Key outcomes point to improved user-provider interaction quality and user knowledge, yet several results reported limited success in changing contraceptive use or long-term contraceptive continuation.

**Table 1:** Synthesis of umbrella review findings on person-centred SRH interventions in primary healthcare settings, globally.

|  | Key interventions and specific tools | Key outcomes | Person-centred domains |
| --- | --- | --- | --- |
| **THEME 1:**<br>**Interactive decision- and counselling support** | **Digital tools to assist users identify family planning preferences, needs, and concerns prior to counselling session (in the waiting room):**<br>Family Planning Quotient and Reproductive Life Index (FPQ/RepLI) tool: incorporated into the electronic medical record.<sup>25 27</sup><br>Smart Choices tool: computer-based tool.<sup>25</sup><br>My Birth Control: tablet-based interactive tool.<sup>25 26</sup><br>MyPath: web-based decision support tool.<sup>25 27</sup><br>Interactive Mobile Application of Contraceptive Choice (iMACC): Mobile application.<sup>25</sup><br>MyFamilyPlan: Web-based preconception health education module.<sup>27</sup><br>**Coaching support:**<br>Smart Patient Coaching: an educator in the waiting room coaches users on how to ask questions and voice their concerns.<sup>26</sup><br>**Counselling techniques:**<br>WHO decision making tool: shared decision-making model allowing providers to share expertise and users to voice their needs, preferences and concerns.<sup>12 26</sup><br>Motivational interviewing: counselling style consisting of collaborative user-provider relationship between using questioning, reflective listening and empathetic statements.<sup>12 26</sup><br>Tailored counselling: adapted counselling based on users' reproductive life stage, social context or prior experience (including emphasis on high-risk populations).<sup>12 27 28 39</sup><br>The Balanced counselling Strategy: providers use job aids to assist in provision of tailored information.<sup>12 26 37</sup><br>WHO Tiered Effectiveness tool: used during counselling to as a visual job aid showing contraceptive methods and effectiveness.<sup>26</sup><br>Checklist to evaluate preconception care needs for HIV patients, which collects information and generates an algorithm, for providers to use.<sup>27</sup><br>Educational videos in clinic waiting rooms about various topics (e.g. condom use, STI prevention) followed by more intensive counselling interventions (one-on-one or group interventions) focused on risky behaviours.<sup>23</sup> | **Positive outcomes:** Digital decision-support tools as well as counselling techniques improved counselling quality and by ensuring preferences were tailored to each user. Digital tools and counselling techniques were successful in increasing knowledge of contraception.<br>**Negative outcomes:** Despite high acceptability, several tools were not successful in changing contraceptive use or long-term continuation. | **Autonomy, dignity, communication, and supportive care** |
| **THEME 2:**<br>**Provider-initiated clinical prompts and support** | **Provider-initiated support:**<br>Proactive provider-initiated HIV testing and counselling for migrants.<sup>36</sup><br>**Electronic medical record (EMR) triggers:**<br>Electronic eligibility reminder for family planning service provision: screening prompt for users in need of family planning<sup>27</sup><br>EMR alerts for contraceptive counselling when users are prescribed teratogenic medications.<sup>31</sup> | **Positive outcomes:** Improved documentation and identification of users needing contraception. EMR prompts reported increased STI screening.<br>**Negative findings:** Some reports of digital prompts hindering clinic | **Communication, and supportive care** |
|  | Automated prompts for STI/HIV screening or testing.<sup>40</sup><br>**EMR tailored documentation:**<br>One Key Question (OKQ): screening tool where provider asks user about pregnancy intention and tailors counselling session accordingly.<sup>25 27</sup><br>Contraceptive vital sign: documentation of pregnancy intentions and contraceptive use by the provider for quick reference.<sup>27</sup><br>Reproductive Life Plan Tool (RLPT): a prompt tool for providers to ask about users' contraceptive needs and refer them accordingly.<sup>27 31</sup> | workflow and speed. While documentation improved record-keeping, there was a gap in recording needs and addressing them. |  |
| **THEME 3:**<br>**Remote and digital information channels** | **Texting:**<br>Two-way interactive SMS reminders for oral and injectable contraceptives or about SRH topics.<sup>29</sup><br>One-way or passive educational text messages and different topics (e.g. SRH, FP, HIV).<sup>29 30 39</sup><br>**Virtual care:**<br>Telemedicine consultations and online prescription platforms.<sup>29</sup><br>Mobile phone-based telemedicine for sexual health care.<sup>21</sup><br>Telephone interviews to provide education on cervical cancer, risks and importance of screening and follow-ups.<sup>34</sup><br>**Media:**<br>Contraception information Tablet App used prior to the clinic visit.<sup>29</sup><br>Interactive smartphone-based game about HIV prevention or mobile phone quiz about reproductive health.<sup>30</sup> | **Positive findings:** Interactive two-way text reminder recorded improvements in contraceptive continuation. Interactive texting and media increased knowledge of SRH topics and improved access for marginalised populations. Virtual care allowed confidential inquiry about sexual health topics.<br>**Negative findings:** non-interactive messages had limited behavioural impact. Some SMS interventions had limited reach due to lack of phone access or digital literacy. | **Privacy & confidentiality, and communication** |
| **THEME 4:**<br>**Integrated service delivery and accessibility strategies** | **Co-location:**<br>Delivering family planning during well-baby visits, within paediatric clinics.<sup>28</sup> or within maternal and child health services to enhance resource efficiency.<sup>32</sup><br>HIV-related educational announcements and invitation for free HIV testing while waiting in the clinic waiting area.<sup>36</sup><br>Brief 'Health talks' about family planning by health assistants before a clinic visit.<sup>39</sup><br>**Accessibility:**<br>Improvement of accessibility through same day LARC insertions.<sup>22</sup><br>Removal of financial barriers and making LARCs free for adolescents.<sup>22</sup><br>Financial incentives for HIV retesting.<sup>33</sup><br>STI self-testing or express testing programs and efficient sample collection services.<sup>40</sup> | **Positive findings:** High user satisfaction for co-location. Increased HIV testing after free invitation to test during regular visits. Increased uptake of LARCs among adolescents. Increased HIV retesting when incentives were introduced.<br>**Negative findings:** requires high facility readiness and administrative coordination. Self-testing initiatives required clearer instructions. | **Health facility environment, dignity, and autonomy** |
| **THEME 5:**<br>**Social and peer-led navigation** | **Navigation:** "Friend to friend" programs and formal patient navigation given by cancer prevention specialists to educate women (especially vulnerable women) about cervical and breast cancer screening, have a discussion about concerns and ask providers for screening.<sup>34 35</sup> | **Positive findings:** Increased knowledge on cancer screening.<br>**Negative findings:** Cultural factors impacted outcomes. | **Trust, dignity, social support and autonomy** |

#### 2. Provider-initiated clinical prompts and support

Interventions in this category depended on health system infrastructure and providers’ initiative to support SRH care (Table 1-Theme 2). The interventions included provider-initiated support, electronic medical records (EMR) prompts, and EMR documentation that specifically collected tailored information about the users, such as pregnancy intentions and contraceptive needs. These interventions generally supported communication and supportive care PCC domains by ensuring reproductive needs were not overlooked. The findings suggested improved clinical documentation, however, some providers reported hinderance in workflow.

#### 3. Remote and digital information channels

This theme described interventions that leveraged technology to provide care. This included mHealth platforms for SMS reminders and educational messages, telemedicine consultations, and media use for educational material (Table 1-Theme3). These modes of delivery were useful in enhancing privacy & confidentiality, and communication PCC domains by allowing private access to sensitive information and overcoming physical and logistical barriers. Positive outcomes included increased knowledge and improved access for marginalized groups, though lack of mobile phone access and digital illiteracy may limit these interventions. Simple one-way text messages were found to be potentially less useful than interactive messaging in ensuring contraceptive continuation.

#### 4. Integrated service delivery and accessibility strategies

This theme represents structure changes to the primary care environment to improve convenience and reduce barriers. Related interventions include co-location of SRH care components during paediatric visits, and strategies to improve accessibility through efficient same day services or remove financial barriers (Table1-Theme4). The interventions addressed the health facility environment, dignity and autonomy PCC domains by creating a responsive environment that respects the user’s time and financial situation. These interventions generally reported higher user satisfaction and increased uptake of effective contraceptive methods, particularly among adolescents.

#### 5. Social and peer-led navigation

This theme focused on human connection and social networks to navigate the health system. The interventions were specifically ‘Friend to friend’ and formal patient navigation for cancer screening (Table1-Theme5). This intervention primarily touched on trust, social support and autonomy domains of PCC, which uses social networks and human connection to build trust and empower individuals to navigate health systems. These interventions can increase knowledge and confidence regarding cervical and breast cancer screening, however, outcomes were sometimes influenced by cultural factors.

### Reporting biases and certainty of evidence

Assessment of risk of bias due to publication or reporting bias was not performed quantitatively by any of the reviews. Six reviews mentioned in their results discussions that their confidence in the evidence was limited either due to lack of study risk of bias assessments, reporting bias or publication bias.

## DISCUSSION

This umbrella review of person-centred SRH interventions in PHC settings identified 21 reviews describing interventions for various SRH topics, mostly related to family planning. These interventions were synthesised and mapped into five themes that focusing on the different modes of delivery within PHCs. These themes were divided into interventions that provided tools for interactive decision- and counselling-support, interventions where providers proactively initiated supportive care, interventions that focused on remote and digital information channels, interventions that provided integrated service delivery and reduced accessibility constraints, and interventions that depended on social and peer-led navigation. The synthesis revealed that SRH interventions often integrate elements of person-centred care, even when not explicitly designed as such. While many interventions reported an improvement in knowledge and uptake, achieving continuation and long-term behavioural change, such as contraceptive use, remained a challenge.

Only four reviews explicitly referred to PCC concepts,^12 25 26 28^ while others described interventions that only implicitly related to a person-centred approach, such as, tailoring information, improving communication and respecting choice. This suggests that while elements of PCC may be included, they are not always intentionally embedded in intervention design. We have inferred person-centredness for most of the interventions in Table 1 and Table S4, yet it is possible that these interventions were performed in a non-PCC manner. Moreover, none of the interventions included all the domains of person-centred care.^5^

While our review focused on identifying person-centred SRH interventions, it is worth noting that there was limited explicit evaluation of person-centred outcomes within the included reviews highlighting a critical gap in current SRH research. There is a need for more robust definition and measurement to ensure holistic care.^9 12 41^ Furthermore, the reviews’ measured outcomes mainly focused on immediate and short-term outcomes.^27 28 31^ There was no follow up of long term impact of person-centred care;^12^ limited research on continued use and support of contraceptive method choice;^25^ and no focus on repeated breast/cervical cancer screening.^35^

Designing SRH interventions through a person-centred lens is crucial for ensuring their quality, cultural appropriateness, and tailoring to different contexts and sub-populations.^12 25 26 34 35^ A review of women’s preferences for contraceptive counselling showed that they valued active engagement with their providers, which includes tailoring counselling to their needs, as well as receiving information about different methods and their side effects, from their providers and through written sources.^42 43^ Successful tailoring includes addressing language barriers,^27^ and overcoming digital illiteracy and communication network issues, which can impact the uptake of digital interventions.^21 29 30^ Furthermore, targeted interventions that account for age, cultural norms, and level of risky behaviour are often more successful.^23^ Culturally tailored approaches have been shown to increase acceptability among immigrant and low-income groups,^34^ with interventions designed for specific ethnicities, culture or languages demonstrating higher engagement.^28 35^ For certain vulnerable groups, such as refugees, SRH interventions in camps proved more accessible than those in dispersed settings,^24^ further emphasising the importance of context-specific design.

The provision of person-centred SRH interventions requires readiness of healthcare settings and providers. Many interventions seemed to address PCC domains, nevertheless, their implementation and acceptability in the PHC setting and among providers requires measurement. Few included reviews measured provider acceptability or clinic workflow impact,^25 27 36^ yet, the results were insufficient to provide insights into barriers. Some eligible reviews referred to provider biases and time constraints as factors influencing interventions impact.^25 26 40^ Ensuring effective implementation of person-centred interventions requires provider training in the provision of PCC and well-established infrastructure and workflow within PHC settings.^5 23 42^

To our knowledge, this is the first umbrella review to integrate multiple areas of SRH person-centred intervention literature. We searched five bibliographic databases to ensure comprehensive coverage. Through this umbrella review, we synthesised a large amount of data and presented useful and practical information. However, the work is not without limitations. Combining multiple topics and review methodologies into a single review led to substantial heterogeneity across the interventions (e.g. different targeted populations or outcome measurements). There was no presentation of intervention effectiveness due to the ‘critically low’ quality of eligible reviews (Table S2). The interventions were described to provide an overview of interventions with the various outcomes documented. However, no comparisons were made due to inconsistent measurements of clinical and/or PCC outcomes. Information on publication and reporting bias was limited. Some reviews noted that restricting their searches to English-language, using a single database or limiting their inclusion to peer reviewed literature may have limited their findings. This suggests that relevant interventions may still be missing from this synthesis. It is also possible that the search missed relevant reviews, despite the detailed search strategy across five databases. It is worth noting that multiple reviews stated that evidence was limited and more research to evaluate intervention effectiveness was required.^22 25 28 29^Additionally, more research is required from LMICs due to the limited geographic scope of current studies.^12 25-28 33 36^

## CONCLUSION

SRH intervention literature was reviewed to identify person-centred SRH interventions implemented in PHCs, globally. The umbrella review highlighted that person-centred SRH interventions were delivered through diverse modes, moving beyond traditional patient-provider clinical care. By categorizing these interventions into five themes this synthesis illustrated a shift towards making SRH care more responsive, accessible and respectful of users, and specifically women’s autonomy. A significant gap remains in the explicit and comprehensive evaluation of person-centredness. Future efforts must prioritize the deliberate integration and rigorous assessment of PCC components to ensure that SRH is not only available as a pillar of universal health coverage but is delivered through supportive models that ultimately respect and respond to the women’s choices, needs and preferences.

## Supporting information

Supplemental materials - detailed methods

## Contributors

GES, SJM, FEK, TKK and SEF planned the umbrella review. SEF, LAJ, PH, and ZK conducted the literature screening, data extraction and quality appraisal with supervision and validation from GES. GES conducted the data synthesis with input from SJM, FEK and TKK. GES wrote the first draft with input from SEF. All co-authors critically reviewed and edited the subsequent drafts of the manuscript. FEK, TKK and SJM were responsible for funding acquisition. SJM and GES are the guarantors of the study.

## Data Availability Statement

All data relevant to the study are included in the article or uploaded as supplementary information.

---

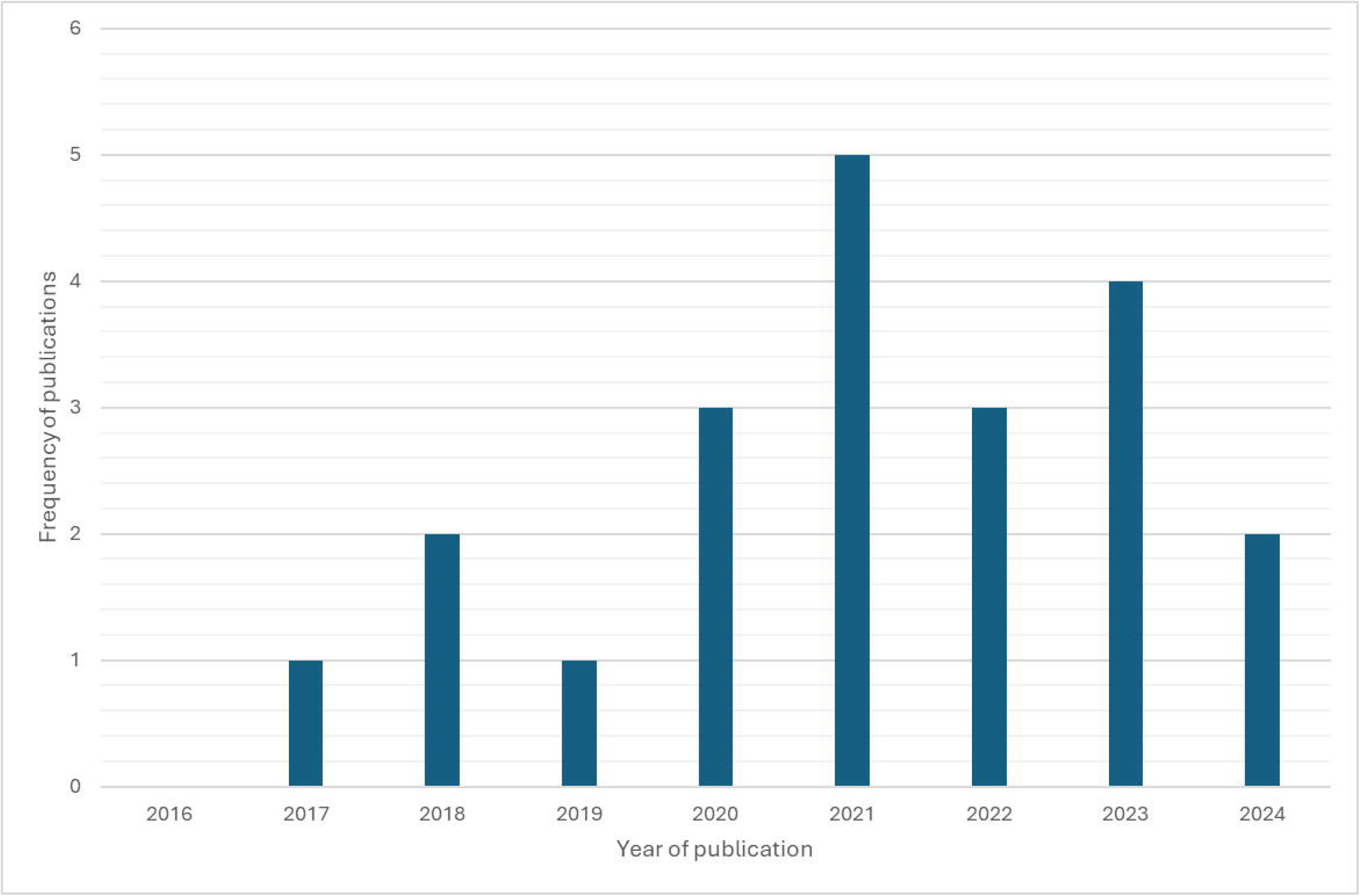

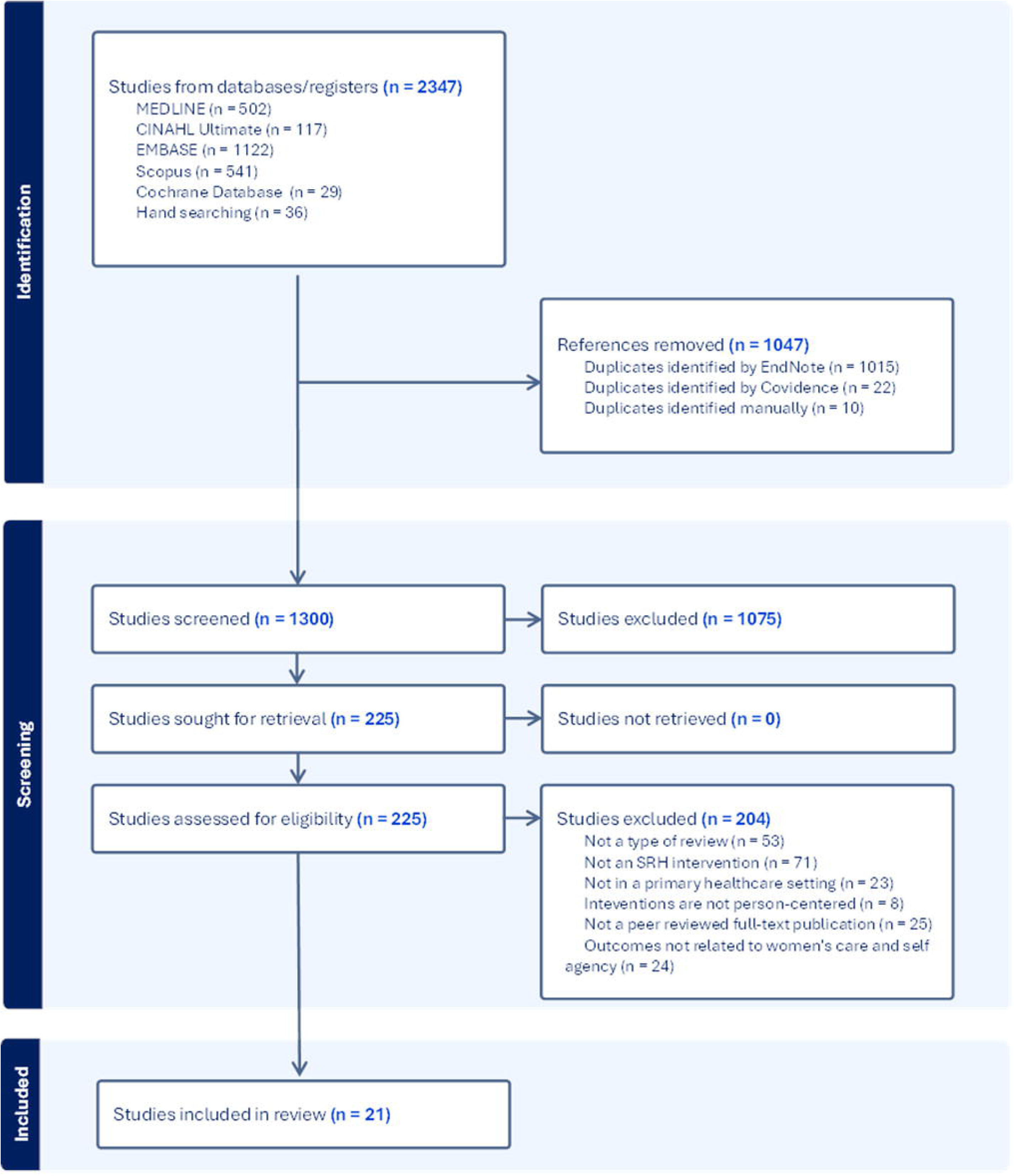

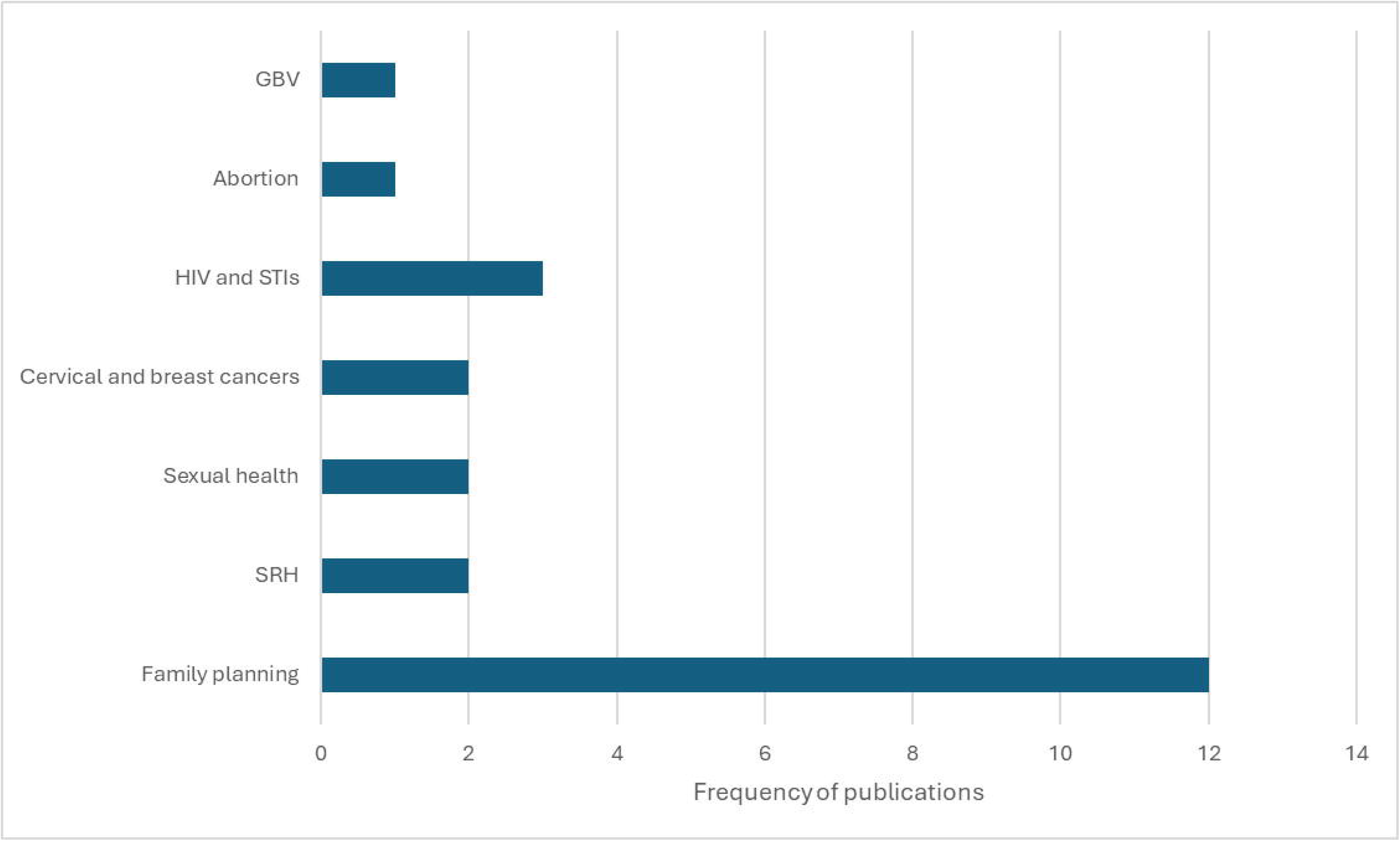

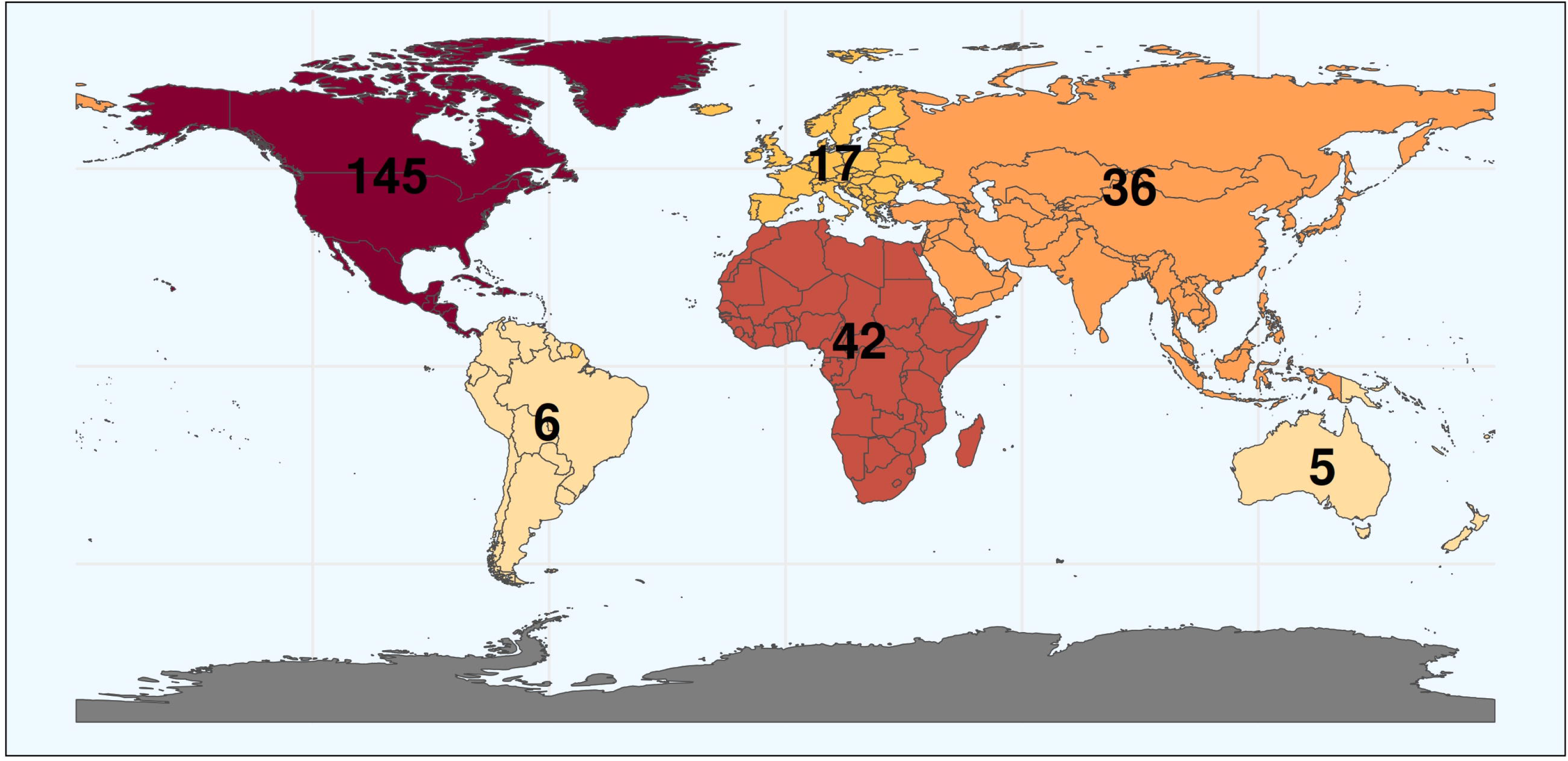

