## Supplemental materials - detailed methods for "Mapping of person-centred sexual and reproductive health interventions at primary healthcare settings: An umbrella review"

**Supplementary Material – DETAILED METHODS**

Supplement to the manuscript title: Mapping of person-centred sexual and reproductive health interventions at primary healthcare settings: An umbrella review

### METHODS

The umbrella review was structured in accordance with the PRIOR (Preferred Reporting Items for Overviews of Reviews) reporting guidelines for overviews of reviews of healthcare interventions.[1] The protocol was registered on Open Science Framework (OSF) Registry, registration DOI: https://doi.org/10.17605/OSF.IO/WC238. Below is a more detailed presentation of the methods following the PRIOR guidelines.

#### Eligibility criteria

The inclusion and exclusion criteria are described using the PICO (Population, Intervention, Comparator, and Outcome) and PICo (Population, Intervention, Context) research question elements. Given the presence of different types of reviews, PICO criteria were used for reviews that assessed intervention outcomes, and PICo characteristics for reviews that only described interventions without comparing them to any other type of intervention or lack of intervention. The inclusion and exclusion criteria were based on the following details.

*Population*: The population of interest was adolescent girls and women, without any specific age range. Boys and men receiving person-centred SRH interventions were included when such interventions led to women's SRH self-agency and autonomy. Reviews that did not include this population were excluded and interventions/primary studies within the reviews that did not fulfil the criteria were not extracted. When there was no disaggregation by gender, the intervention was included.

*Intervention*: Person-centred SRH interventions delivered in PHC settings were included in the umbrella review. Person-centred care was based on Sudhinaraset et al (2017)'s definition for person-centred reproductive health care, which consists of eight domains: dignity, autonomy, privacy & confidentiality, communication, social support, supportive care, trust and health facility environment.[2] When not explicitly stated, person-centredness of SRH interventions were inferred from one or more of these domains. Other terms included and were synonymous with person-centred care were people-centred care, patient-centred care, women-centred care, user-centred care, client-centred care, individual-centred care, holistic care, comprehensive care.

Interventions, programs, approaches, and digital innovations were included in the following SRH areas: family planning; contraception provision and counselling; sexually transmitted infections (STI) prevention/screening; HIV screening/vaccination; menstrual health; menopause management; adolescent SRH; cervical cancer screening/prevention; human papillomavirus (HPV) screening/vaccination; sexual health and wellbeing; comprehensive sex/sexuality education; and sexual health counselling. Interventions that focused on obstetric care and pregnancy/childbirth related care were excluded. Furthermore, interventions that were solely social media-based, community-based, home-based or tertiary hospital-based interventions were excluded. Interventions in eligible reviews that did not align with an person-centred domains or PHC setting were not extracted.

*Comparison*: For reviews that assessed intervention outcomes, any comparator was considered. Reviews with no comparator also were considered if they presented person-centred SRH interventions.

*Context*: To be included, reviews had to describe SRH interventions that were implemented within PHC settings; interventions offered in the community, schools, homes or tertiary hospital settings, or were social media-based were excluded. All geographic locations were included.

*Outcomes*: Included outcomes ranged from clinical outcomes such as increased uptake and continuation of care, to PCC outcomes, such as, self-agency, autonomy, increased satisfaction, service user/provider interaction and communication, mental well-being, increased knowledge and perception. This list of outcomes is not exhaustive and was kept broad to inlcude outcomes that each eligible review defined. Outcomes related to intervention cost evaluations were excluded.

*Additional elements:* Any type of review was eligible for inclusion. The time frame of interest was from 2016 to present. In 2016, the World Health Assembly convened and strongly supported the resolution to strengthen health services through implementation of integrated people-centred health services.[3 4] No language restrictions were applied.

**Information Sources**

On September 15, 2024, we searched five electronic bibliographic databases: MEDLINE (Ovid), CINAHL Ultimate (EBSCOhost), EMBASE (Elsevier), Scopus (Elsevier) and Cochrane Database of Systematic Reviews (Cochrane Library).

**Search Strategy**

A comprehensive search strategy was developed, receiving input from specialists in SRH research and a Medical Librarian. Search terms were identified based on existing literature that would be eligible for this umbrella review. The strategy focused on four concepts: SRH topic areas, intervention terms, PHC context and reviews. Search terms for PCC were not included because the term and its synonyms are not widely used, hence this would have limited the search results. A comprehensive list of Medical Subject Headings (MeSH) controlled vocabulary (if applicable in the database) and keywords was used for each concept to ensure capturing the highest number of eligible reviews. Table S1 shows the search strategy for Ovid MEDLINE consisting of MeSH controlled vocabulary and keyword searches. Similar MeSH and keyword terms were used for Elsevier EMBASE and EBSCOhost

CINAHL databases. For Elsevier Scopus and Cochrane Library of Systematic Reviews, the searches were limited to keywords (TableS1).

**Selection Process**

The literature search results were exported into Endnote and duplicates removed. The results were then exported into Covidence Systematic Review software where more duplicates were removed, and the software was used to managing the screening and data extraction processes. Inclusion and exclusion screening criteria were developed for the title/abstract and full-text screening stages, which were entered into Covidence software. Prior to beginning the screening process, four researchers (SEF, LAJ, PH, ZK) conducted a calibration exercise to pilot the screening criteria and discussed inconsistencies until there was consensus.

Subsequently, in pairs, the researchers independently screened titles/abstracts of the results. In case of disagreements, the researchers met to come to a resolution. When necessary, a third researcher (GES) was consulted to make the final decision. Following this screening, the four researchers (SEF, LAJ, PH, ZK), in pairs, independently screened the full-text articles for inclusion in the umbrella review and recorded the reason for exclusion. The researchers met to reach a consensus whenever there were disagreements and in case it was not resolved, a third researcher (GES) was consulted and/or the primary studies of disagreed upon interventions were reviewed. Furthermore, given the aim of the umbrella review was to describe the existing relevant interventions, there was no exclusion of primary studies or interventions that overlapped between the eligible reviews.

**Data collection process**

A data extraction form was developed and uploaded to Covidence software. The form included sections on the review aims, methods, and research elements. Four researchers (SEF, LAJ, PH, ZK) tested the data extraction form and discussed inconsistencies and ambiguous fields until there was clarity and consistency in extraction. Data was extracted from each eligible review, independently by two researchers (SEF & LAJ or PH & ZK). Any discrepancies were resolved through discussion, and a third researcher (GES) was consulted when needed. A third reviewer (GES) validated the final extracted data.

The standardized data extraction form includes information about the publication (author, publication year, citation, objectives/aims of the review), the review methodology (including information on type of review, population/s included, setting and geographic scope, number of databases searched, date range of search) and the synthesis of results (number of eligible studies included in the review, SRH topics covered, description of intervention components, explicit/implicit PCC domains, factors about intervention implementation, review limitations/recommendations).

To identify the degree of overlap between primary studies in eligible reviews, a citation matrix was created listing the relevant primary studies in eligible reviews. Subsequently, the degree of overlap was measured using the corrected covered area (CCA) formula and the overlapping primary studies were assessed.[5 6]

**Data Items**

All clinical and person-centred outcomes (defined above) that were presented in the synthesised results of the eligible reviews were included since this umbrella review aimed to map and describe the relevant person-centred SRH interventions. The effect measures were reported as presented in eligible reviews, even if heterogeneous. The reported outcome measurements included odds ratios, prevalence ratios, proportions, or narrative presentation.

**Risk of Bias Assessment**

The quality of eligible reviews was assessed using the AMSTAR 2 critical appraisal tool for systematic reviews, which includes randomized or non-randomized studies of healthcare interventions.[7] The checklist was implemented on Covidence software and the four researchers (SEF, LAF, PH, ZK), in pairs, independently assessed the eligible reviews. The overall rating and confidence in the review results was calculated based on the type of review and whether it satisfies the checklist's critical and non-critical criteria, as described by Shea et al (2017).[7]

The quality of primary studies in the reviews was assessed based on information collected from the eligible reviews on the risk of bias. Lack of quality assessments were documented, and no re-assessment was made since the aim of the umbrella review is to describe the existing evidence.

**Synthesis Methods**

Extracted data was synthesised narratively. This was planned given the high heterogeneity in interventions and outcome measures in the eligible reviews. Moreover, since all types of reviews were included, meta-analysis was not possible. In the narrative synthesis, reviews and interventions were grouped based on the topics and type of interventions. The overall positive and negative outcomes for each group of interventions were explored and the person-centred domains that best addressed the interventions were identified by the researchers.

**Reporting Bias Assessment and Certainty Assessment**

To document risk of bias due to missing results arising from reporting biases, one researcher (GES), collected related information from the eligible reviews, and subsequently, summarized the findings and potential gaps. Certainty assessment for outcomes of interest was also collected by the researcher (GES) from the eligible reviews when present. For both assessments, there was no re-assessment conducted given the primary aim of the umbrella review was to identify existing interventions rather than assess effectiveness.
