## Supplemental materials - Tables for "Mapping of person-centred sexual and reproductive health interventions at primary healthcare settings: An umbrella review"

**Supplemental Material - TABLES**

Supplement to the manuscript titled: Mapping of person-centred sexual and reproductive health interventions at primary healthcare settings: An umbrella review

**Table S1:** OVID Medline Search Strategy for umbrella review on person-centred SRH interventions in primary healthcare settings

| Concept | Search terms |
|---|---|
| **SRH topics** | MeSH Terms<br>reproductive health/ or sexual health/ or women's health/ or exp Contraception / or exp Sexually Transmitted Diseases/ or Menopause/ or Menstruation/ or Menstrual cycle/ or Papillomavirus Vaccines/ or sexuality/ |
| | Keyword terms<br>("reproductive health" or "((sex*) adj2 (health or wellbeing or well-being))" or "((women or woman) adj2 health)" or "family plan*" or contracept* or sexuality or "sexually transmitted disease prevention" or "sexually transmitted disease screening" or "sexually transmitted infection prevention" or "sexually transmitted infection screening" or menopaus* or menstrua* or "papillomavirus vaccin*" or "HIV vaccin*" or "cervical cancer screening" or "cervical cancer prevention").ti,ab,kw. |
| **Intervention terms** | MeSH Terms<br>exp health education/ or delivery of health care/ or digital health/ or exp counseling/ or Psychosocial Intervention/ or implementation science/or health services/ |
| | Keyword terms<br>(interven* or program* or campaign* or approach* or counsel* or promot* or educat* or service*).ti,ab,kw. |
| **Primary health care settings** | MeSH Terms<br>exp Primary health care/ or Community Health Centers/ or community mental health centers/ or Maternal-Child Health Center/ or General Practice/ or Family practice/ |
| | Keyword terms<br>(((communit* or neighbor* or neighbour*) adj2 health adj2 (center* or centre* or clinic or clinics)) or ((maternal or woman or women) adj2 health adj2 (center* or centre* or clinic or clinics)) or (outpatient adj2 (clinic or clinics)) or ((primary or "first line") adj2 (healthcare or "health care" or "medical care")) or ((general or family) adj2 practice*)).ti,ab,kw. |
| **Review type** | MeSH Terms<br>"review literature as topic"/ or systematic reviews as topic/ |
| | Keyword terms<br>(review or meta-analysis or "meta analysis").ti,ab,kw. |

**Table S2:** Description of the 21 eligible reviews in the umbrella review on person-centred SRH interventions in primary healthcare settings, globally, 2016-2024

| First author (year) | Title of the review | Aim of the review | Type of review with number of studies included | Geographic scope of included studies | Search strategy bibliographic databases, supplementary searches (timeframe in years) | Targeted population(s) | Findings synthesis |
|---|---|---|---|---|---|---|---|
| Meekers et al. (2024) | Tools for patient-centred family planning counselling: A scoping review | To identify key tools to make family planning care more patient-centred, to review the domains of patient-centred care they address, and to identify gaps in the evidence base | Scoping review of 33 studies<br><br>Eligible studies for this review: 13 | North America (n=11)<br>Africa (n=1)<br>Asia (n=1) | PubMed, SCOPUS<br>(2013-2022) | All women | Narrative |
| Diamond-Smith et al. (2018) | Interventions to improve the person-centred quality of family planning services: a narrative review | To describe interventions related to person-centred care and identify their effect on service users' experiences and family planning outcomes such as knowledge, uptake, and continuation | Narrative review of 25 studies | North America & Europe (n=11)<br>Africa (n=5)<br>Asia (n=5)<br>South America (n=3) | PubMed, CINAHL, EconLit.<br>Supplementary searches: grey literature (dissertations, theses, government reports, non-governmental organization reports,<br>and funder reports), relevant journals, organization websites, contacted key personnel in relevant organizations<br>(1990-2015) | General population | Narrative |
| Danna et al. (2021) | Leveraging the Client-Provider Interaction to Address Contraceptive Discontinuation: A Scoping Review of the Evidence That Links Them | To summarize global literature on contraceptive counselling approaches, focusing on identifying techniques and tools that enhance client-provider interactions to reduce method discontinuation. And to examine counselling approaches tailored to | Scoping review of 54 studies.<br><br>Eligible studies for this review: 11 studies | North America (n=8)<br>South America (n=2)<br>Asia (n=2) | PubMed, EMBASE, PsycINFO.<br>Supplementary searches: Google Scholar and Google grey literature searches<br>(1990-2018) | All women, and adolescents and young adults (24 years and younger) | Narrative |

| | | | | | | | |
|---|---|---|---|---|---|---|---|
| | | adolescents, who face additional barriers. | | | | | |
| Ren et al. (2023) | Preconception, Interconception, and reproductive health screening tools: A systematic review | To identify and describe the standardized inter-conception and preconception screening tools for reproductive health needs that are applicable in general outpatient clinical practice | Systematic review of 53 studies<br><br>Eligible studies for this review: 29 studies | North America (n=29) | PubMed, Web of Science, CINAHL<br>(2000-2022) | All women | Narrative |
| Karlin et al. (2024) | A Scoping Review of Patient-Centered Perinatal Contraceptive Counseling | To synthesize research on patient-centred perinatal contraceptive counselling, focusing on patient preferences, experiences, and health outcomes to propose best practices and identify research gaps | Scoping review of 34 studies<br><br>Eligible studies for this review: 19 studies | North America (n=11)<br>Europe (n=6)<br>Oceania (n=2) | PubMed, Embase, Popline<br>(Popline: 1992-2019<br>PubMed, Embase: 1992-2022) | Reproductive aged service user receiving care in a clinical setting | Narrative |

| | | | | | | | |
|---|---|---|---|---|---|---|---|
| Thompson et al. (2020) | Telemedicine for family planning: A scoping review | To identify and synthesize evidence on the use of telemedicine for family planning | Scoping review of 43 studies<br><br>Eligible studies for this review: 14 studies | North America (n=12)<br>Asia (n=2) | Cochrane Library, Cochrane Collaboration Registry of Controlled Trials, EMBASE, PubMed, MEDLINE (2008-2019) | All population seeking contraception and medication-abortion services | Narrative |
| Feroz et al. (2021) | Using mobile phones to improve young people sexual and reproductive health in low- and middle-income countries: A systematic review to identify barriers, facilitators, and range of mHealth solutions | To identify mHealth solutions that can be used for improving young people's SRH in LMICs and highlight facilitators and barriers for adopting mHealth interventions | Systematic review of 15 studies | Africa (n=13)<br>Asia (n=2) | PubMed, CINAHL Plus, Science Direct platform, Cochrane Central. Supplementary searches: grey literature (non-published, internal or non-reviewed papers, repositories) (2005-2020) | Adolescents and youth (10–24 years old). | Narrative |

| | | | | | | | |
|---|---|---|---|---|---|---|---|
| Shatilwe et al. (2021) | Evidence on access to healthcare information by women of reproductive age in low- and middle-income countries: Scoping review | To evaluate the accessibility to healthcare information for women of reproductive age in LMICs | Scoping review of 4 studies<br><br>Eligible studies for this review: 1 study | Asia (n=1) | Science Direct, PubMed, EBSCOhost, CINAHL, MEDLINE, PsycINFO, Emerald, Embase, CDSR. Supplementary searches: grey literature (Google Scholar, organizational projects, reference lists of included studies, conferences and websites. (2004-2020) | Women aged 14-49 years old | Narrative |
| Burgess et al. (2018) | A systematic review of the effect of reproductive intention screening in primary care settings on reproductive health outcomes | To assess the effect of reproductive intention screening in primary care on reproductive health outcomes | Systematic review of 9 studies | North America (n=6)<br>Europe (n= 3) | Ovid MEDLINE, PubMed, CINAHL, Embase, CDR/DARE, Web of Science, ISRCTN registry, ClinicalTrials.gov, Cochrane Library.<br>Supplementary searches: hand search and contacted individuals for additional resources (expert consultations).<br>(2000-2017) | Patients of reproductive age (15–49 years old). | Narrative |

| | | | | | | | |
|---|---|---|---|---|---|---|---|
| Wilkes et al. (2020) | Use of Long-Acting Reversible Contraceptives Amongst Adolescents: An Integrative Review | To identify strategies that increase the use of LARCs among adolescent populations | Integrative Review of 15 studies | North America (n=15) | CINAHL, MEDLINE, Embase. Supplementary search: backward search of included articles references. (2007-2016) | Adolescents aged 14 to 19 years old | Narrative |
| Duminy et al. (2021) | Urban Family Planning in Low- and Middle-Income Countries: A Critical Scoping Review | To present the key dimensions and challenges of urban growth in LMICs, offer a critical review of recent research findings on urban family planning and fertility dynamics, and highlight priorities for future research | Scoping review of 279 studies.<br><br>Eligible studies for this review: 6 study | Africa (n=5)<br>Asia (n=1) | Web of Science. Supplementary searches: Google Scholar, review of references in identified articles, contacted key experts in the field. (2000-2020) | Women of reproductive age in urban settings | Narrative |

| | | | | | | | |
|---|---|---|---|---|---|---|---|
| Akwara et al. (2023) | The Urban Environment and Disparities in Sexual and Reproductive Health Outcomes in the Global South: a Scoping Review | To explore the drivers, barriers and contextual factors related to SRHR in urban areas in the Global South, and to describe interventions in urban areas to improve SRHR outcomes | Scoping review of 115 studies.<br><br>Eligible for this review: 11 studies | Africa (n=11) | PubMed, MEDLINE, SCOPUS, COCHRANE Library, Web of Science, POPLINE, JSTOR. Supplementary searches: Google scholar, review of refences in identified articles<br>(2010-2022) | General urban populations | Narrative |
| Onaisi et al. (2022) | Sexual risk behaviour reduction interventions in primary care in Organization of Economic Cooperation and Development countries. A systematic review | To analyse sexual risk behaviours reduction interventions proposed in primary care in the Organization of Economic Development and Cooperation (OECD) countries | Systematic review of 30 studies | North America (n=28)<br>Europe (n=1)<br>Oceania (n=1) | PubMed, Cochrane Library, EBSCOhost (PsycArticle, Psychology and Behavioural Sciences Collection, PsycINFO and SocINDEX), Scopus, CAIRN. Supplementary searches: Google Scholar, review of references in identified systematic reviews.<br>(from database inception – 2021) | All populations | Narrative |

| | | | | | | | |
|---|---|---|---|---|---|---|---|
| Mariño et al. (2023) | Educational interventions for cervical cancer prevention: a scoping review | To identify, map, and describe characteristics of educational interventions for cervical cancer prevention in adult women | Scoping review of 33 studies.<br><br>Eligible for this review = 22 studies | North America (n=7)<br>South America (n=1)<br>Asia (n=11)<br>Africa (n=2)<br>Europe (n=1) | CINAHL, MEDLINE PubMed, SciELO, PsycINFO, Cochrane Library database, LILACS, and Nursing Database (BDENF) database, Web of Science, Embase, Scopus.<br>Supplementary searches: Google Scholar,<br>Catalog of Theses and Dissertations CAPES - Brazil and Brazilian<br>Registry of Clinical Trials – ReBEC, Digital Library of Theses and Dissertations (BDTD)<br>(time frame not reported) | Adult women | Narrative |
| Atere-Roberts et al. (2020) | Interventions to increase breast and cervical cancer screening uptake among rural women: a scoping review | To review and assess the published literature on interventions to increase BCC screening in rural communities | Scoping review of 8 studies<br><br>Eligible for this review = 3 studies | North America (n=3) | PubMed<br>Supplementary searches: review of references in identified articles.<br>(2006-2019) | Women residing in rural communities | Narrative |

| | | | | | | | |
|---|---|---|---|---|---|---|---|
| Aung et al. (2 17) | Interventions for Increasing HIV Testing Uptake in Migrants: A Systematic Review of Evidence | To review and evaluate interventions that aim to increase HIV testing uptake in migrant populations | Systematic review of 10 studies<br><br>Eligible for this review = 3 studies | North America (n=1)<br>Europe (n=1)<br>Africa (n=1) | PubMed, Web of Science, Embase, CINAHL, PsycInfo. Supplementary searches: Google Scholar, Google. (1985-2016) | Migrants aged 15 years or older | Narrative |
| Sorhaindo et al. (2022) | Interventions to reduce stigma related to contraception and abortion: a scoping review | To identify the types, volume and characteristics of available evidence and analyse gaps in the knowledge base for interventions to reduce contraception and abortion stigma | Scoping review of 18 studies.<br><br>Eligible for this review = 2 studies | Asia (n=1)<br>North America (n=1) | MEDLINE, PubMed, Embase, Web of Science PsycINFO.<br>Supplementary searches: grey literature search in websites of key organizations.<br>(2000-2022) | All women | Narrative |

| | | | | | | | |
|---|---|---|---|---|---|---|---|
| AlHamawi et al. (2023) | Family planning interventions in Jordan: A scoping review | To describe family planning interventions implemented in Jordan and highlight gaps | Scoping review of 10 studies.<br><br>Eligible for this review = 4 studies | Asia (n=10) | PubMed. Supplementary searches: Review of references in identified articles. (2010-2022) | All populations | Narrative |
| Boydell et al. (2022) | Getting intentional about intention to use: A scoping review of person-centred measures of demand. Studies in Family Planning | To synthesis knowledge about intention to use (ITU) contraception and related measurements | Scoping review of 112 studies.<br><br>Eligible for this review = 3 | Africa (n=2)<br>Asia (n=1) | PubMed, Web of Science. (1990-2020) | All women | Narrative |

| | | | | | | | |
|---|---|---|---|---|---|---|---|
| Footman et al. (2021) | A Systematic Review of New Approaches to Sexually Transmitted Infection Screening Framed in the Capability, Opportunity, Motivation, and Behaviour Model of Implementation Science | To summarize work that has evaluated clinical implementation of chlamydia and gonorrhoea screening programs and adoption of new technologies | Systematic review of 14 studies | North America (n=8)<br>Europe (n=4)<br>Oceania (n=2)<br>Africa (n=1) | PubMed, Embase, CINAHL, Scopus, Cochrane Library, HSRProj.<br>(2015-2020) | All populations | Thematic analysis based on the Capability, Opportunity, Motivation, and Behavior (COM-B) model |
| Munyuzangabo et al. (2019) | Delivery of sexual and reproductive health interventions in conflict settings: a systematic review | To synthesize the available literature on SRH intervention delivery in conflict settings to inform potential priorities for further research and additional guidance development | Systematic review of 110 studies | Africa (n=83)<br>Asia (n=32)<br>Europe (n=2)<br>South America (n=5) | MEDLINE, Embase, CINAHL, and PsycINFO.<br>Supplementary searches: grey literature search in 10 major humanitarian agencies and organizations.<br>(1990-2018) | Conflict-affected populations and internally displaced people | Narrative |

SRH: Sexual and reproductive health; SRHR: Sexual and reproductive health rights; LMICs: Low- and middle-income countries; LARC: Long acting reversible contraceptives; BCC: breast and cervical cancer; ITU: Intention to use.

**Table S3:** Quality assessment of eligible reviews about person-centred SRH interventions based on AMSTAR 2 Quality Appraisal tool

| Author, Year | 1. Did the research questions and inclusion criteria for the review include the components of PICO? | 2. Did the report of the review contain an explicit statement that the review methods were established prior to the conduct of the review and did the report justify any significant deviations from the | 3. Did the review authors explain their selection of the study designs for inclusion in the review? | 4. Did the review authors use a comprehensive literature search strategy? | 5. Did the review authors perform study selection in duplicate? | 6. Did the review authors perform data extraction in duplicate? | 7. Did the review authors provide a list of excluded studies and justify the exclusions? | 8. Did the review authors describe the included studies in adequate detail? | 9.1. Did the review authors use a satisfactory technique for assessing the risk of bias (RoB) in individual studies that were included in the review? | 9.2. Did the review authors use a satisfactory technique for assessing the risk of bias (RoB) in individual studies that were included in the review? | 10. Did the review authors report on the sources of funding for the studies included in the review? | 11. If meta-analysis was performed did the review authors use appropriate methods for statistical combination of results? | 12. If meta-analysis was performed, did the review authors assess the potential impact of RoB in individual studies on the results of the meta-analysis or other evidence synthesis? | 13. Did the review authors account for RoB in individual studies when interpreting/ discussing the results of the review? | 14. Did the review authors provide a satisfactory explanation for, and discussion of, any heterogeneity observed in the results of the | 15. If they performed quantitative synthesis did the review authors carry out an adequate investigation of publication bias (small study bias) and discuss its likely impact on the results of the review? | 16. Did the review authors report any potential sources of conflict of interest, including any funding they received for conducting the | Overall confidence in the results of the review |
|---|---|---|---|---|---|---|---|---|---|---|---|---|---|---|---|---|---|---|
| **Meekers , 2024** | Yes | No | Yes | Partial yes | Yes | Yes | No | Partial yes | For RCTs: No | For NRSIs: No | No | No meta-analysis | No meta-analysis | No | No | No meta-analysis | Yes | **Critically low** |
| **Diamond-Smith , 2018** | Yes | Yes | No | Partial yes | No | No | No | Partial yes | For RCTs: Partial yes | For NRSIs: Partial yes | No | No meta-analysis | No meta-analysis | Yes | Yes | No meta-analysis | Yes | **Low** |
| **Danna , 2021** | Yes | No | Yes | Partial yes | No | No | No | No | For RCTs: No | For NRSIs: No | No | No meta-analysis | No meta-analysis | No | No | No meta-analysis | Yes | **Critically low** |
| **Ren , 2023** | Yes | No | Yes | Partial yes | Yes | No | No | Partial yes | For RCTs: Partial yes | For NRSIs: Partial yes | No | No meta-analysis | No meta-analysis | No | No | No meta-analysis | Yes | **Critically low** |
| **Karlin , 2024** | Yes | Yes | No | Partial yes | Yes | Yes | No | Partial yes | For RCTs: No | For NRSIs: No | No | No meta-analysis | No meta-analysis | No | No | No meta-analysis | Yes | **Critically low** |
| **ThompsonT-A , 2020** | Yes | Partial yes | Yes | Partial yes | No | No | No | No | For RCTs: No | For NRSIs: No | No | No meta-analysis | No meta-analysis | No | No | No meta-analysis | Yes | **Critically low** |
| **Feroz , 2021** | Yes | Yes | Yes | Partial yes | Yes | Yes | No | Partial yes | For RCTs: Partial yes | For NRSIs: Partial yes | No | No meta-analysis | No meta-analysis | Yes | Yes | No meta-analysis | Yes | **Low** |
| **Shatilwe , 2021** | Yes | Partial yes | Yes | Partial yes | Yes | Yes | Yes | Partial yes | For RCTs: Partial yes | For NRSIs: Partial yes | No | No meta-analysis | No meta-analysis | Yes | No | No meta-analysis | Yes | **Moderate** |
| **Burgess , 2018** | Yes | Partial yes | No | Yes | Yes | Yes | No | Partial yes | For RCTs: Partial yes | For NRSIs: Partial yes | No | No meta-analysis | No meta-analysis | Yes | No | No meta-analysis | Yes | **Low** |
| **Wilkes , 2020** | Yes | No | No | Partial yes | No | No | No | No | For RCTs: Partial yes | For NRSIs: Partial yes | No | No meta-analysis | No meta-analysis | No | No | No meta-analysis | Yes | **Critically low** |
| **Duminy , 2021** | Yes | No | Yes | Partial yes | No | No | No | Partial yes | For RCTs: No | For NRSIs: No | No | No meta-analysis | No meta-analysis | No | Yes | No meta-analysis | Yes | **Critically low** |
| **Akwara , 2023** | Yes | No | Yes | Partial yes | No | No | No | Partial yes | For RCTs: No | For NRSIs: No | No | No meta-analysis | No meta-analysis | No | No | No meta-analysis | Yes | **Critically low** |

| | | | | | | | | | | | | | | | | | | |
|---|---|---|---|---|---|---|---|---|---|---|---|---|---|---|---|---|---|---|
| **Onaisi , 2022** | Yes | No | Yes | Partial yes | Yes | No | No | Partial yes | For RCTs: Partial Yes | For NRSIs: Partial yes | No | No meta-analysis | No meta-analysis | Yes | Yes | No meta-analysis | Yes | **Critically low** |
| **Mariño , 2023** | Yes | No | Yes | Partial yes | Yes | No | No | Partial yes | For RCTs: No | For NRSIs: No | No | No meta-analysis | No meta-analysis | No | No | No meta-analysis | Yes | **Critically low** |
| **Atere-Roberts , 2020** | Yes | No | No | No | Yes | No | No | Partial yes | For RCTs: No | For NRSIs: No | No | No meta-analysis | No meta-analysis | No | Yes | No meta-analysis | No | **Critically low** |
| **Aung , 2017** | Yes | No | No | Partial yes | No | Yes | No | Partial yes | For RCTs: Partial yes | For NRSIs: Partial yes | No | No meta-analysis | No meta-analysis | No | No | No meta-analysis | Yes | **Critically low** |
| **Sorhaindo , 2022** | Yes | No | Yes | Partial yes | Yes | No | No | Partial yes | For RCTs: No | For NRSIs: No | No | No meta-analysis | No meta-analysis | No | No | No meta-analysis | Yes | **Critically low** |
| **AlHamawi , 2023** | Yes | No | Yes | No | No | No | No | Partial yes | For RCTs: No | For NRSIs: No | No | No meta-analysis | No meta-analysis | No | No | No meta-analysis | Yes | **Critically low** |
| **Boydell , 2022** | Yes | No | No | Partial yes | No | No | No | Partial yes | For RCTs: No | For NRSIs: No | No | No meta-analysis | No meta-analysis | No | No | No meta-analysis | Yes | **Critically low** |
| **Footman , 2021** | Yes | No | Yes | Partial yes | Yes | No | No | Partial yes | For RCTs: No | For NRSIs: No | No | No meta-analysis | No meta-analysis | No | No | No meta-analysis | Yes | **Critically low** |
| **Munyuzangabo , 2020** | Yes | Partial yes | Yes | Partial yes | No | No | No | Partial yes | For RCTs: No | For NRSIs: No | No | No meta-analysis | No meta-analysis | No | Yes | No meta-analysis | Yes | **Critically low** |

Note: AMSTAR 2 critical domains are items 2, 4, 7, 9, 11, 13, 15.

**Table S4:** Intervention characteristics and main results of the eligible reviews in the umbrella review on person-centred SRH interventions in primary healthcare settings, globally, 2016-2024

| **First author (year)**<br>**Title** | **Types of interventions presented in the review** | **Potential Person-Centred Care domain** | **Overall findings by outcome and factors that may influence interventions (if mentioned)** | **Review limitations/ identified evidence gaps/recommendations** |
|---|---|---|---|---|
| Meekers et al. (2024) [1]<br><br>Tools for patient-centred family planning counselling: A scoping review | **SRH topics targeted in these interventions:** interventions and tools to make family planning counselling more patient-centred.<br><br>**Description of interventions in PHCs:**<br>**a)** One Key Question (OKQ) is a screening tool, where the provider asks the service user: *Would you like to become pregnant in the next year? (responses: Yes/No/Unsure/ I'm fine either way)*. Subsequently, the provider chooses the patient-centred counselling that is applicable to the provided response. The counselling includes preconception counselling for those who want to get pregnant and contraception methods counselling for those who don't want to. Service users who responded 'don't know' or 'fine either way' were offered both counselling content.<br>**b)** Family Planning Quotient and Reproductive Life Index (FPQ/RepLI) is composed of 4 components to assist in discussions and decision making about reproductive and family planning goals. The 1st component is a visual graph in the service user's medical record showing the reproductive plan (completed with the help of a health educator before consulting the provider. The 2nd part contains a decision-making tree for contraceptive method choices. The 3rd part is a tool to track progress and relevant outcomes. The 4th component is a table the tracks yearly changes in contraception.<br>**c)** Smart Choices is an electronic tool composed of a questionnaire completed by service users in the waiting room. The provider receives the responses and is able to understand the service user's concerns/needs/ preferences and provide more tailored counselling. The second component of the tool contains an interactive | **Intervention a:** Autonomy/ dignity/ communication/ supportive care<br>**Intervention b:** Autonomy/ dignity/ communication/ supportive care<br>**Intervention c:** Autonomy/ dignity/ communication/ privacy & confidentiality<br>**Intervention d:** Autonomy/ dignity/ communication/ privacy & confidentiality<br>**Intervention e:** Autonomy/ dignity/ communication/ privacy & confidentiality/ supportive care<br>**Intervention f:** Autonomy/ dignity/ communication/ privacy & confidentiality/ social support | • **Ease of use/acceptability**: A comparison of tool (a) and (b) among service users found that service users were more likely to find tool (b) to be more useful to track reproductive goals (51% for (a) vs 76% for (b), p-value 0.02). A post-test for tool (b) found that 92% of service users thought it was helpful and would use it, while 91% of providers reported it was helpful for reproductive health discussion and 83% said it was needed. A qualitative assessment of (d) among providers found that the tool was feasible to incorporate and acceptable to service users, while some stated it hindered clinic flow and was possibly difficult for service users not used to tech. For (e), most service users reported that the tool was easy to understand and comfortable to answer. A qualitative assessment of (f) found that providers and service users said tool was self-explanatory and easy to use, and providers said it reduced their workload. Time for completion was: 5min for (b), 15min for (c), 11min for (e), 15min for (f).<br>• **Quality of care and provider-service user interaction**: A comparison of tool (a) and (b) among service users and providers showed that service users found both helpful to communicate their preferences and goals to the providers. With (a), documentation of reproductive plan decreased pre- vs post (22% vs 6%, p-value=0.02), documentation of counselling and contraceptive method remained the same. For tool (b), over 90% of service users said it helped them think about and communicate their reproductive goals, and over 90% of providers said the tool was | **Limitations:**<br>• Limited to two databases<br>• Language bias due to search restriction to English language only<br>• Most studies focused on initial contraceptive method choice, with limited attention to post-adoption counselling or ongoing support<br>• Evidence of counselling tools effectiveness is limited.<br>• Conclusions about relative effectiveness of tools were not possible because the review did not do bias/quality assessments.<br><br>**Evidence gaps:**<br>• Limited regional representation<br>• Evidence of the effectiveness of these tools requires more research.<br>• Lack of studies on provider resistance to adopting tools, their effect on workload, and patient flow.<br><br>**Recommendations:** |

| | | | | |
|---|---|---|---|---|
| | guide that provides service users with information about all contraceptive methods.<br>**d)** My Birth Control is a tablet-based interactive tool for family planning decision-making. The tool is completed by the service user before consulting the provider. The tool gives information about contraceptive methods, asks service users about their preferences, medical history to help recommend a contraceptive method accordingly.<br>**e)** MyPath is a web-based tool for family planning decision-making to facilitate reproductive health counselling in primary care visits. The tool is used prior to seeing the provider. It collects information about the service user's feelings and preferences about pregnancy and childbearing. It also provides information about menstruation, fertility pre-pregnancy health and suitable contraceptive methods.<br>**f)** Interactive Mobile Application for Contraceptive Choice (iMACC) is a mobile phone intervention for postpartum women to assist in family planning decision making and facilitate the counselling sessions. The tool is self-administered in the clinic waiting room prior to seeing the provider and can be used as a stand-alone tool for women. The tool asks about health history, service user preferences and concerns, as well as partner's attitude towards family planning. | | helpful to understand service user preferences and focus counselling, while 73% said family planning counselling improved. tool (c) was more patient-centred compared to control group (score 3.9 vs. 3.7, p-value<0.05) and sexual health topics were discussed more (1.2 vs 0.9, p-value<0.10). For tool (d), interpersonal counselling quality increased (OR=1.45;95%CI:1.03-2.05), satisfaction with side-effect information increased (OR=1.61; 95%CI:1.11-2.33), no effect on service user satisfaction with provider assistance of contraceptive choice, but providers found tool (d) increased efficiency of the session by allocating time to what service user wants to discuss. For (e), tool made counselling more efficient for providers and service users discussed pregnancy or contraceptive needs more often (93% intervention group vs. 68% control group, p-value<0.05), intervention service users had higher self-efficacy in communication scores (0.8 vs. 0.2, p-value<0.05), but no improvement in communication quality. For tool (f), service users were satisfied with its confidentiality.<br>• **Contraceptive knowledge**: Post-test of tool (c) found that service users knew about more contraceptive methods compared to control group (11.1 vs. 10.7, p-value<0.001). Tool (d) improved contraceptive knowledge where intervention group more likely to know that IUDs are more effective than pills (OR = 2.65;95%CI:1.94–3.62), methods that stop period are safe (OR=1.86;95%CI:1.28–2.71),and implants do not affect fertility (OR=1.54;95%CI:1.14–2.07). Tool (e) increased knowledge scores compared to control group (1.7 vs. 0.2, p-value<0.01). Tool (f) assisted service users in awareness of contraceptive options and possible side-effects.<br>• **Contraceptive decision-making, use and continuation**: Tool (a) did not change in | • Studies need to use standardized indicators<br>• More studies on provider workload and understanding the effect of changing the counselling technique to the effect of using the tool.<br>• There is a need for expansion of the evidence base on acceptability and effectiveness through systematic reviews and meta-analyses.<br>• More research is required to assess whether improvements in experience due to the tools leads to better family planning outcomes. |

| | | | |
|---|---|---|---|
| | | | current contraceptive method. Tool (c) led to intervention service users to be less likely to choose IUDs/implants (9% vs. 20%) instead of oral contraceptive (64% vs. 54%, p-value<0.10), compared to control group. Tool (d) did not record significant effect on satisfaction of chosen method or continuation after 7 months, yet providers reported service users being more engaged in method selection. Tool (e) showed no significant change in likelihood of changing from non-prescription to prescription methods. Tool (f) empowered service users to make informed decisions about their preference method.<br><br>**Factors that may influence interventions:**<br>i. Time constraints to proper implementation of interventions<br>ii. Provider biases<br>iii. Availability of clinic resources |

| | | | | |
|---|---|---|---|---|
| Diamond-Smith et al. (2018) [2]<br><br>Interventions to improve the person-centred quality of family planning services: a narrative review | **SRH topics targeted in interventions:** family planning use/uptake, continuation, or knowledge Interventions, as well as person-centred care (satisfaction, measures of quality of care, service user/provider interactions, changes in self-efficacy or power).<br><br>**Description of interventions in PHCs, by component:**<br>**a)** Communication interventions (11 studies)- information about topics related to FP, RH and STIs broadly or those that provided tailored and person centred FP knowledge and counselling.<br>**b)** Privacy/confidentiality interventions (6 studies) - information given to be read privately, e.g. handouts and leaflets, audiovisual tools or DVDs, interactive tools like mobile-phone application and computer assisted interventions<br>**c)** Dignity interventions - emphasized high risk populations on broader societal/ cultural factors (e.g. empowering women re gender/power dynamics, discussing GBV, sexual and power negotiation, resisting peer pressure, dealing with psychosocial and relationship factors and tailored counselling with PCC approach.<br>**d)** Autonomy interventions - focused on improving decision making power in family planning choice. E.g.: helping clients write down questions for the provider and practice them, self-efficacy and effective use of methods, guidance to identify needs, decision-making support, helping service users remember methods, find accessible methods, build negotiation power.<br>**e)** Social support interventions - focused on increasing social support among partners/family members in decisions about family planning. E.g. one intervention engaged partners alone, another focused on couples, and the third one focused on encouraging youth to involve parents. | Interventions in this review were explicitly described based on person-centred domains. The mentioned domains are: Communication/ privacy & confidentiality/ Dignity/ autonomy/ social support | • **Family planning initiation/uptake:** 5/11 communication interventions (a) had a positive impact and the rest no impact on uptake. 2/4 privacy interventions (b) showed a positive effect. 5/11 dignity interventions (c) reported a positive effect on uptake. 0/5 autonomy interventions (d) had any impact. 1/2 social support interventions had a positive impact on uptake (e).<br><br>• **Family planning continuation:** 3/6 communication interventions (a) reported lower discontinuation. 0/3 privacy interventions (b) showed any impact on continuation. 2/4 dignity interventions (c) has a positive effect on continuation. 0/3 autonomy interventions had any impact on continuation (d). 2/2 social support interventions had a positive impact (e).<br><br>• **Person-centred outcomes:** 11 unique studies measured these outcomes. 7/8 communication interventions (a) reported improvement in this outcome. 1/1 privacy intervention (b) has a positive impact. 4/4 dignity interventions (c) showed improvement. 5/5 autonomy interventions (d) had a positive impact. 1/2 social support interventions (e) had a positive effect on these outcomes.<br><br>• **Family planning knowledge:** 6/7 communication interventions (a) increased knowledge. 3/3 privacy interventions (b) had a positive impact. 4/5 of dignity interventions (c) had a positive effect on knowledge. 2/3 autonomy interventions (d) had a positive impact. | **Limitations:**<br>• Limited comparability due to variability in study designs, focus, methodology, and outcome measures.<br>• Most studies were conducted in the developed world, with limited evidence about what works in developing countries.<br>• There were challenges among the research team to define interventions based on person-centred care frameworks.<br>• Only included English-language studies<br>• Risk of bias was assessed but there was no additional quality assessments or exclusion of studies based on initial risk of bias.<br><br>**Evidence gaps:**<br>• The evidence from multi-component interventions was limited due to poor quality of relevant research on these interventions<br><br>**Recommendations:**<br>• There is a need for more thorough quantitative studies to measure person-centred outcomes.<br>• Mixed methodologies and qualitative data are required to identify new |

| | | | | |
|---|---|---|---|---|
| | | | | measures of person-centred quality of care.<br>• Future designs of person-centred care interventions need to be sensitive to cultural norms in different contexts.<br>• More evidence is necessary to understand how family planning knowledge and quality of interventions can lead to behaviour change in the short and long term.<br>• Future interventions should consider including more than one domain of person-centred care.<br>• Studies should research the long-term impacts of person-centred care processes and health behaviours such as unintended pregnancies, abortions, and fertility rate. |

| | | | | |
|---|---|---|---|---|
| Danna et al. (2021) [3]<br><br>Leveraging the Client-Provider Interaction to Address Contraceptive Discontinuation: A Scoping Review of the Evidence That Links Them | **SRH topics targeted in these interventions:** Contraceptive counselling interventions.<br><br>**Description of interventions in PHCs:**<br>**a)** WHO decision-making tool for Family planning service users and providers: consists of a shared decision making model allowing providers to share technical expertise and clients to voice their needs, preferences and concerns.<br>**b)** The Balanced Counselling Strategy: tool that uses job aid to assist in provision of tailored information to service users.<br>**c)** Smart Patient Coaching: is an intervention where an educator is present in the clinic waiting room to coach service users regarding how to ask questions, how to talk about concerns, how to ask for clarifications from providers.<br>**d)** Motivational Interviewing: is a patient-centred counselling style which consists of creating a collaborative relationship between service users and provider, using questioning, reflective listening, empathetic statements in the aim of identifying the intrinsic motivation for clients' behaviour changes.<br>**e)** My Birth Control: a tablet-based tool provided to clients before consulting the provider that assists clients choose their contraceptive method based on preferences and values.<br>**f)** The WHO Tiered Effectiveness Tool: it is a visual job aid that shows the contraceptive methods and their effectiveness for pregnancy prevention on a spectrum. | **Intervention a:** Dignity/ autonomy/ communication<br>**Intervention b:** autonomy/ communication<br>**Intervention c**: Autonomy/ dignity/ communication/ supportive care<br>**Intervention d:** Autonomy/ dignity/ communication/ supportive care<br>**Intervention e:** Autonomy/ communication<br>**Intervention f:** Autonomy/ communication | • **Quality of care & client-provider interactions:** Intervention (a) improved counselling by increasing service user engagement and making the sessions more tailored to their needs, as well as improving the exchange of information by acting as a job aid for providers and a decision aid for service users. Intervention (b) increased contraceptive use among service users. Intervention (c) enhanced service user engagement with the provider. Intervention (d) doubled the initiation and continuation of LARCs among women aged 15–29 after 3 months. Intervention (e) led to higher rating on quality, informed decision-making and knowledge. Intervention (f) was no evaluated.<br><br>• **Contraceptive discontinuation:** Interventions (a), (b), and (e) had no impact on discontinuation. Intervention (c) recorded lightly reduced discontinuation rates (3.9% vs. 7.8%) at 8 months.<br><br>**Factors that may influence interventions:**<br>• Barriers: Provider biases, time constraints, and insufficient training.<br>• Enablers: Service user engagement, comprehensive counselling on side effects, and tailored decision-making aids. | **Limitations:**<br>• Included studies varied in research methodologies and definitions of the outcome of interest.<br>• The review did not assess the quality of the evidence, because it was out of scope.<br><br>**Evidence gaps:**<br>• Many of the findings were from mid- to high-income countries, with limited studies in Sub-Saharan Africa.<br>• There are limited studies that evaluate interventions for their impact on discontinuation of contraceptives.<br>• There is minimal evidence that existing tools/trainings improve the interaction between service user and provider and improve health outcomes.<br><br>**Recommendations:**<br>• There is a need for rigorous evaluation of counselling approaches to identify their impact on outcomes.<br>• Further research on person-centred counselling approaches in various contexts is necessary. |

| | | | | |
|---|---|---|---|---|
| Ren et al. (2023) [4]<br><br>Preconception, Interconception, and reproductive health screening tools: A systematic review | **SRH topics targeted in these interventions:** Preconception and reproductive health screening tools.<br>**Description of interventions in PHCs:Specific tools:a)** Contraceptive vital sign: health record documentation of women's pregnancy intentions and contraceptive use for providers' quick reference.<br>**b)** Family Planning Quotient (FPQ) and Reproductive Life Index (RefLI): Visual tool representing women's reproductive goals to be used by providers for goal-oriented care.<br>**c)** Gabby: web-based virtual health counsellor<br>**d)** MyFamilyPlan: web-based preconception health education module<br>**e)** MyPath: Web-based decision support tool that consists of reproductive goals review, information about having optimal health for pregnancy and contraceptive decision support.<br>**f)** One Key Question (OKQ): Physicians ask service users one question: "Do you intend to become pregnant in the next year", in order to provide appropriate counselling.<br>**g)** READY-Girls: preconception counselling for teenage girls with type1 diabetes. The program includes watching 2 CDs, reading a book and having a brief counselling session with a nurse during 3 visits over a period of 9 months.<br>**h)** Reproductive Health Self-Assessment Tool (RH-SAT): Survey on pregnancy which includes information on reproductive health topics.<br>**i)** Reproductive Life Plan Tool (RLPT): Prompt for providers to ask women about contraceptive needs and offer referral services when contraceptive services are required.<br>**Standardized approaches:**<br>**j)** Checklist to evaluate preconception care needs of HIV patients: checklist consists of 2 pages that collect patient information and generates an algorithm for providers to use.<br>**k)** Preconception risk assessment and brief counselling: Questionnaire for assessing reproductive health risk followed by counselling for 10-15 minutes. | **Intervention a:** Communication/ supportive care.<br>**Intervention b:** Autonomy/ dignity/ communication/ supportive care.<br>**Intervention c:** Autonomy/ communication/ privacy & confidentiality.<br>**Intervention d:** Autonomy/ communication/ privacy & confidentiality.<br>**Intervention e:** Autonomy/ dignity/ communication/ privacy & confidentiality/ supportive care.<br>**Intervention f:** Autonomy/ communication/ supportive care.<br>**Intervention g:** Autonomy communication/ privacy & confidentiality/ supportive care<br>**Intervention h:** Autonomy/ communication/ supportive care<br>**Intervention i:** Autonomy/ communication/ supportive care | • **Provision of quality contraceptive services/counselling:** Intervention (a) led to better information documented about contraceptive use, however, no evidence of improvement in care. Intervention (b) assisted in service user-provider communication and decision-making process; it also provided contraception education and acted as a family and reproductive goals planning tool. Intervention (e) led to an improvement in quality of reproductive decisions while not overwhelmed providers' workload. Intervention (f) was reported to increase the rate of preconception screening. Intervention (h) can potentially improve service user-provider communication on reproductive health. Intervention (k) may be suitable to identify low-income women needing reproductive health services. Intervention (l) resulted in an increase of family planning use. Intervention (n) increased reproductive health knowledge and helped women with chronic diseases make more informed choices<br>.• **SRH behaviours:** Intervention (c) was associated with decrease in preconception risk. Intervention (d) led to significantly higher proportion of women talking about reproductive health with their providers. Intervention (e) led to an increase in the proportion of primary care visits where reproductive needs were discussed. Intervention (g) potentially has long term impacts on teenagers' knowledge, intentions and beliefs regarding seeking health providers to discuss reproductive health behaviours and outcomes. Intervention (h) was reported to assist women in discussing contraception with their providers. Intervention (m) led to a reduction in the risk of rapid subsequent adolescent births.<br>• **Acceptability and feasibility:** Intervention (c) results showed that it can be feasible | **Limitations:**<br>• Only included studies in English and conducted in USA.<br>•The review excluded studies that focused on contraceptive use (including method selection, continuation, etc.), unless the tool was clearly applicable to preconception counselling.<br><br>**Evidence gaps:**<br>• Lack of long-term follow-up studies on service user outcomes.<br>• Insufficient evidence on specific outcomes such as pregnancy timing and health behaviour change<br><br>**Recommendations:**<br>• Longitudinal study designs are required to ideally track service users' multiple outcomes. |

| | | | |
|---|---|---|---|
| | **Electronic Health Record additions.**<br>**l)** Electronic eligibility reminder for family planning services provision: electronic screening prompt that identifies people in need of family planning<br>**Motivational Interviewing:**<br>**m)** Computer assisted motivational interviewing (CAMI): CAMI algorithms provide summary about teens being low, medium or high risk for pregnancy and STIs based on their responses to intentions about current sexual relations and contraceptive and condom use. Subsequently interventionists do a 20min motivational interviewing session to motivate teenagers to use contraceptives.<br>**Reproductive life plans questionnaires**<br>**n)** Reproductive life plan for women with chronic diseases: a long survey that assists women in setting goals according to their pre-existing pregnancy risk factors and chronic conditions | **Intervention j:** dignity/ communication/ supportive care<br>**Intervention k:** dignity/ communication/ supportive care<br>**Intervention l:** Communication/ supportive care<br>**Intervention m:** Dignity/ communication/ supportive care<br>**Intervention n:** Autonomy/ dignity/ communication/ supportive care | medium for ongoing counselling care for adolescents and young adults. Intervention (e) was accepted by women. Intervention (f) did not affect the clinic workflow. Intervention (i) was deemed feasible in paediatric clinics during routine infant care by the providers. Intervention (j) was acceptable in assisting conversations of reproductive intentions. Intervention (l) was reported to be accepted and feasible in a health centre setting. |

| | | | | |
|---|---|---|---|---|
| Karlin et al. (2024) [5]<br><br>A Scoping Review of Patient-Centred Perinatal Contraceptive Counselling | **SRH topics targeted in these interventions:** Perinatal contraceptive counselling interventions.<br><br>**Description of interventions in PHCs:**<br>**a**) Antenatal contraceptive counselling and provision of postpartum contraception by midwives<br>**b)** Immediate postpartum counselling with written educational material<br>**c)** Toolkit-based intervention for immediate postpartum LARC counselling and provision<br>**d)** Offering participants written literature or an educational video.<br>**e)** Co-locating contraceptive services with well-baby visits.<br>**f)** Standard postpartum counselling plus supplemental "holistic" in-person counselling at 35 weeks of pregnancy with printed and online written material, in addition to a SMS reminder at week 37).<br>**g**) Expert contraceptive care offered antenatally by specialized family planning nurses.<br>**h)** Postpartum counselling in addition to incorporating information on healthy birth spacing and LARC methods. | **Interventions a:** Autonomy/ Communication<br>**Interventions b:** Autonomy/ Communication/ privacy& confidentiality<br>**Interventions c:** Autonomy/ Communication<br>**Interventions d**: Autonomy/ privacy & confidentiality<br>**Intervention e:** Autonomy/ communication supportive care<br>**Intervention f:** Autonomy/ communication/ / privacy & confidentiality/ supportive care<br>**Intervention g:** Autonomy/ dignity/ communication<br>**Intervention h:** Autonomy/ Communication/ privacy& confidentiality | • **Structure, process and timing of perinatal contraceptive counselling:** Adolescent participants in intervention (a) reported that timing of counselling at 22 weeks was 'about right' (81%) and it was 'very or quite helpful' (81%).<br><br>• **Association of perinatal counselling with service user experience:** Intervention (b) had no impact on satisfaction between the intervention and control groups, yet those in the intervention group were more likely to report that written material assisted in ultimate decision for contraceptive method compared to the control group (p-value<0.01). Intervention (c) recorded poor service user satisfaction. Comparing intervention (d) with provider-service user counselling found that service user satisfaction was generally high across three groups yet there was a trend to increased satisfaction among provider counselling group (p-value<0.05) compared to other groups. Intervention (e) recorded high satisfaction rates (80%) and 64% of participants said they recommend linking both content, while those who did not accept the linked visit stated they didn't want a new provider, or didn't want to discuss contraception with their children, or did want the consultation to prolong. Intervention (f) participants had higher satisfaction with contraceptive experience. Participants who received intervention (g) found counselling at the antenatal period "helpful".<br><br>• **Association of perinatal counselling with health outcomes (rapid repeat pregnancy)**: Intervention (h) did not show any difference in pregnancy rates at 6 and 12 months, compared to those who received only counselling. Intervention (g) did not have any association with repeat pregnancy rates at 1 year period. | **Limitations**:<br>• Many articles were excluded due to our focus on specific patient-centred outcomes.<br>• The findings were focused on United States, Canada, Europe, the UK, New Zealand, and Australia only.<br>• The review did not include articles published in non-English languages.<br>• The quality of data synthesis is as good as the extracted findings from studies<br><br>**Evidence gaps:**<br>• There is a lack of longitudinal studies assessing long-term health outcomes.<br>• There is limited focus on and disaggregation by diverse populations, particularly racial/ethnic minorities.<br>• No study explicitly considers transgender or trans-expansive service users, and no included studies focused on cis-gender females.<br><br>**Recommendations**:<br>• Future research should collect data on service user preferences of care as well as health outcomes. |

| | | | | |
|---|---|---|---|---|
| Thompson et al. (2020) [6]<br><br>Telemedicine for family planning: A scoping review | **SRH topics targeted in the interventions:** Telemedicine provision for contraceptive behaviours<br><br>**Description of interventions in PHCs:**<br>**a)** 3-month mobile phone-based intervention with 6 automated interactive messages and counsellor phone support when needed<br>**b)** Tablet computer with a contraception information app to be used for up to 15min, in addition to standard care<br>**c)** Educational text message interventions<br>**d)** Daily interactive text message reminders on oral and injectable contraceptives continuation<br>**e)** Daily text message reminders on oral contraceptive pill adherence<br>**f)** Online platforms for hormonal contraceptive prescriptions through remote health assessments and video consultations | **Intervention a:** Autonomy/ privacy & confidentiality/ communication/ Supportive care<br>**Intervention b:** Autonomy/ communication/ health facility environment<br>**Intervention c:** Privacy & confidentiality/ Communication<br>**Intervention d:** Communication/ supportive care<br>**Intervention e:** Communication/ supportive care<br>**Intervention f:** Dignity/ autonomy/ privacy & confidentiality/ communication/ Supportive care | • **Contraceptive knowledge:** Significant increase in contraceptive knowledge among service users who received intervention (b). Interventions in (c) had limited positive effect on increasing contraceptive knowledge.<br>• **Contraceptive initiation/uptake:** Intervention (a) led to increase in uptake of effective contraception methods. Intervention (b) recorded increase in interest for implants. Intervention (c) did not improve uptake.<br>• **Contraceptive continuation:** Intervention (d) recorded higher continuation rates in one study, higher proportion of service users returning for follow-up appointments on another study and 3 times higher odds of continuing effective contraception after the intervention in a third study (OR:3.65, 95%CI:1.26-10.08) compared to no reminders.<br>• **Adherence to oral contraceptives:** Intervention (e) had no effect on use, missed pills or refills. Reminders to fulfil advanced emergency contraceptive prescriptions had a limited additive effect after each message.<br>• **Provision of evidence-based contraceptive care:** Intervention (f) varied in modalities and processes, yet overall, platforms screened potential contraceptive users appropriately.<br><br>**Factors that may influence interventions:**<br>• Funding for telemedicine initiation<br>• Provider literacy in computers and eHealth<br>• Connectivity and technology infrastructure<br>• Reimbursement challenges from the insurance companies because of limited awareness of these services.<br>• Restrictions related to the existing family planning services e.g. service user security/privacy. | **Limitations**:<br>• Captures peer-reviewed studies of interventions only.<br>• Restricted to English language only.<br>• There is no critical appraisal of the included studies.<br><br>**Evidence gaps:**<br>• Lack of evidence on SRH telemedicine applications that evaluate how knowledge will impact contraceptive use and continuation or how it will lead to better counselling practices by providers.<br>• Many SRH applications do not provide accurate or comprehensive contraceptive information. And it is not known if they are developed by experts and use medically credible information.<br>• There is an absence of regulations for telemedicine applications making their incorporation in healthcare settings difficult.<br><br>**Recommendations:**<br>• Provision of contraceptives through telemedicine requires further research |

| | | | | |
|---|---|---|---|---|
| Feroz et al. (2021) [7]<br><br>Using mobile phones to improve young people sexual and reproductive health in low- and middle-income countries: A systematic review to identify barriers, facilitators, and range of mHealth solutions | **SRH topics targeted in these interventions**: mHealth for SRH education and behaviour change communication<br><br>**Description of interventions in PHCs:**<br>**a)** m4RH project is a text-based system for improving family planning information access<br>**b)** short messaging service on SRH information<br>**c)** Text to change - text messages for HIV/AIDS campaign<br>**d)** 6001 service - passive texting service for sexual health information<br>**e)** Unidirectional (SMS messages of reproductive health information) and interactive text messaging (reproductive health quiz game with financial incentives).<br>**f)** Interactive mobile phone quiz for reproductive health (with incentives)<br>**g)** Interactive smartphone-based game related to HIV prevention | **Intervention a:** Privacy & confidentiality/ Communication<br>**Intervention b**: Privacy & confidentiality/ Communication<br>**Intervention c:** Privacy & confidentiality/ Communication<br>**Intervention d:** Privacy & confidentiality/ Communication<br>**Intervention e:** Autonomy/ Privacy & confidentiality/ Communication<br>**Intervention f:** Dignity/ Autonomy/ Privacy & confidentiality/ Communication<br>**Intervention g:** Dignity/ autonomy/ privacy & confidentiality/ Communication | • **Access to SRH knowledge:** Intervention (a) led to high levels of reach and accessibility of family planning information. Intervention (b) and (g) showed effectiveness in delivering educational programs on HIV prevention. Interventions (b), (e) & (f) led to better reproductive health knowledge among teenagers. One version of intervention (b) assessed SRH knowledge pre- and post-intervention and found improvement (OR=2.7, 95%CI:2.47-2.94 vs OR=3.4, 95%CI:2.99-3.81). Intervention (c) had limited impact in increasing HIV/AIDS knowledge because the SMS platform offered information only to those who answered correctly. Additionally, one evaluation of intervention (b) and (c) showed negative outcomes because of restrictions on telephone use. Intervention (e) was assessed at 3 and 15 months and showed that there was improvement in reproductive health knowledge, that persisted at 15 months. Interventions (b) & (d) showed improvement in sexual health knowledge and safer sexual behaviours.<br>• **SRH outcomes:** Intervention (a) recorded changes in usage of family planning by users, with users reporting improved contraception use. Intervention (g) resulted in better retention of service users for HIV care and ART adherence. Intervention (f) found that mHealth interventions can reach marginalized populations at greater risk of poor SRH outcomes.<br>• **Perception of clients using mHealth:** Intervention (g) was strongly accepted by targeted adolescents and further content was requested by participants.<br><br>**Factors that may influence interventions:**<br>• Barriers: Low digital literacy, network coverage issues, limited linguistic abilities, cost of services and socio-cultural norms that may not mHealth interventions. | **Limitations**:<br>• The interventions and outcomes were heterogenous across the studies and made it difficult to conduct a meta-analysis.<br>• The studies did not adopt a consistent taxonomy to explain the mHealth applications.<br>• Many studies combined multiple mHealth interventions, impeding unique assessment of each intervention.<br><br>**Evidence gaps:**<br>• While mobile phone-based education and behaviour change communication showed strong evidence, other applications like sensors, point-of-care diagnostics, registries/vital events tracking and decision support remain underexplored.<br><br>**Recommendations:**<br>• More evidence is required to comprehensively understand the impact of these interventions on young people's SRH.<br>• Clearer understanding is required regarding the barriers of using mHealth interventions. |

| | | | | |
|---|---|---|---|---|
| | | | • Enablers: Privacy and confidentiality, ease of use, and monetary incentives. | |
| Shatilwe at al. (2021) [8]<br><br>Evidence on access to healthcare information by women of reproductive age in low- and middle-income countries: Scoping review | **SRH topics targeted in these interventions**: Access to healthcare information.<br><br>**Description of intervention in PHCs:**<br>**a)** Use of telemedicine to improve access to healthcare services through video conference telemedicine and mobile phone-based telemedicine. | **Intervention a:** Autonomy/ dignity/ privacy & confidentiality/ communication | • **Connectivity to healthcare information and knowledge:** Intervention (a) has a positive effect of travel restrictions, treatment expenses and apprehension about provider consultations. The mobile phone intervention allowed women and girls to ask confidentially about sexual health and without fear or shyness.<br><br>**Factors that may influence interventions:**<br>• Owning a mobile phone was necessary<br>• Digital literacy | **Limitations**:<br>• The inclusion and exclusion criteria of the review may have been too narrow.<br>• The review excludes studies that were conducted before 2004.<br><br>**Evidence gaps:**<br>• Lack of research on how to address challenges related to digital literacy and lack of access to mobile phones.<br>• Limited published literature about strategies for enabling women to access healthcare information generally, in LMICs.<br><br>**Recommendations:** |

| | | | | |
|---|---|---|---|---|
| | | | | • Future pilot studies and randomized control trials to evaluate strategies that help women access health information.<br>• Further research on interventions to access quality of care and health outcomes. |
| Burgess et al. (2018) [9]<br><br>A systematic review of the effect of reproductive intention screening in primary care settings on reproductive health outcomes | **SRH topics targeted in these interventions:** Reproductive intention screening interventions<br><br>**Description of interventions in PHCs:**<br>**a)** Service user-provider semi-structured discussions on a 'Reproductive Life Plan' adapted to women with chronic diseases such as hypertension, obesity and/or diabetes (RLPC).<br>**b)** Semi-structured discussions on a 'Reproductive Life Plan' with a comprehensive educational component focusing on family planning or folate use depending on women's intentions.<br>**c)** Electronic health system-based screening - three types: 1- digital intake forms to identify reproductive intentions; 2-patient kiosks containing an interactive computer program with information on contraceptives and ability to ask for prescription; 3- electronic medical record with a clinical decision support tool that alerts providers about contraceptive counselling when women are prescribed teratogenic medications.<br>d**)** Screening question on reproductive intentions in a questionnaire for eligibility for contraceptive counselling and referral to counselling if eligible.' | **Intervention a:** Autonomy/ dignity/ communication/ supportive care<br>**Intervention b:** Autonomy/ communication/ supportive care<br>**Intervention c:** Autonomy/ privacy and confidentiality/ supportive care**Intervention d:** Autonomy/ communication/ supportive care | • **Contraceptive use:** Intervention (a) had no effect on contraceptive use. Intervention (e) increased contraception use at three months (OR 1.4, 95%CI:0.3-5.7), though not significantly. Intervention (f) showed that women in need of contraceptive counselling and received it were 2.7 (95%CI:1.5-4.9) times more likely to report using hormonal contraception use and 2.2 (95%CI:0.8-6.5) times more likely to use a highly effective method at last intercourse compared to women in need who did not get counselling.<br>• **Provision of contraceptive services:** Intervention (c) had no effect on provision of new contraceptive services among physicians, yet documentation of contraception in service user records increased significantly between intervention and control group (77% [95%CI:70.7-84.1] in intervention compared to 3% [95%CI:1.5-5.0] in control group).<br>• **Reproductive knowledge:** Intervention (b) reported a statistically significant increase in knowledge on reproduction and folic acid supplementation (from score 6.4 to 9.0) compared to control.<br>• **Perception of clients:** Intervention (b) had a positive experience on 90% of the participants. Intervention (c) reported 85-95% satisfaction with the computer module, yet 65% reported they preferred discussing contraception with a provider; other modes of intervention (c) were highly acceptable by participants. Intervention (d) had a 92.3% | **Limitations:**<br>• The review search strategy included many imprecise terms making the search broader.<br><br>**Evidence gaps:**<br>• Lack of studies and assessments on long term outcomes of reproductive intentions interventions.<br><br>**Recommendations:**<br>• Further research is needed to assess the effectiveness of these interventions on outcomes to inform healthcare practices and wide scale implementation.<br>• Future research needs to assess outcomes both on the short term and long-term health effects. |

| | | | | |
|---|---|---|---|---|
| | | | satisfaction rate with counselling services received.<br><br>**Factors that may influence interventions:**<br>• Socio-economic disparities | |
| Wilkes et al. (2020) [10]<br><br>Use of Long-Acting Reversible Contraceptives Amongst Adolescents: An Integrative Review | **SRH topics targeted in the interventions:** of Long-Acting Reversible Contraceptives use among adolescents<br><br>**Description of interventions in PHCs:**<br>**a)** Strategies related to making LARCs free of charge and accessible to adolescents.<br>**b)** Strategies to improve care accessibility and convenience to service users at clinics, included same-day LARC insertions (The CHOICE Project)<br>**c)** Counselling and education on LARCs. | **Intervention a:** Dignity/ autonomy<br>**Intervention b:** Dignity/ autonomy/ supportive care<br>**Intervention c:** Dignity/ Autonomy/ Communication | • **Contraceptive - LARCs- accessibility:** Intervention (a) led to a substantial increase in adolescents choosing LARCs over less effective methods. One study reported that 69% of adolescents selected LARCs when the price barrier was eliminated. Intervention (b) led to increased uptake of LARCs.<br>• **Contraceptive -LARCs- knowledge:** Intervention (c) increased service user education on LARCs among adolescents.<br><br>**Factors that may influence interventions:**<br>• National policies concerning LARCs.<br>• Types of providers capable of offering LARCs.<br>• Cost of LARCs<br>• Provider training on provision and insertion of LARCs. | **Limitations:**<br>• Six out of the 15 selected articles were on one intervention (CHOICE).<br>• While the review aim was to provide evidence for nurse practitioners in Canada about LARCs, none of the reviewed studies took place in Canada.<br>• The age range was restricted to adolescents only.<br>• Parental consent to choosing LARCs was not considered<br><br>**Evidence gaps:**<br>• There is limited evidence of the use of |

| | | | | |
|---|---|---|---|---|
| | | | | LARCs among adolescent populations.<br><br>**Recommendations:**<br>• Training of nurse practitioners to prescribe and insert LARCs can increase access especially in rural and remote communities. |
| Duminy et al. (2021) [11]<br><br>Urban Family Planning in Low- and Middle-Income Countries: A Critical Scoping Review | **SRH topics targeted in the interventions:**<br>Family planning and contraceptive use interventions<br><br>**Description of interventions in PHCs:**<br>**a)** The Urban Reproductive Health Initiative (URHI), succeeded by The Challenge Initiative (TCI), are large-scale programs that comprised of supply-side (quality and availability of services) and demand-side components (community outreach activities, television and radio programs) to increase contraceptive use among urban populations. They were implemented in Nigeria, Senegal, Kenya and India.<br>**b)** Reproductive health programs that focus on family planning integration with maternal and child health services for enhanced resource efficiency. | **Intervention a:** Supportive care/ Health facility environment.<br>**Intervention b:** Supportive care | • **Contraceptive use:** All country URHI programs were positively impactful. In Kenya, modern contraceptive use among urban women increased from 45% to 52%. In Nigeria, use increased from 21% to 31%.<br>• **Contraceptive initiation/uptake:** Across the African countries with intervention (a), the number of new users attending facilities increased. Yet offering educational information at facilities increased the number of users attending facilities in Nigeria but not in Kenya or Senegal.<br>• **Unintended pregnancies**: Evaluations of intervention (b) suggest a significant opportunity to decrease unintended pregnancies in urban settings.<br>**Factors that may influence interventions:**<br>• social norms surrounding the use of contraception<br>• Communication with spouse<br>• Provider time and workload constraints<br>• Social, economic and political context | **Limitations**<br>• Limited number of studies on urban family planning, which was broad and varied in aim, methodology, geographic scope and themes.<br><br>**Evidence gaps:**<br>• Research concerning the influence of social norms on contraceptive use decision-making is needed.<br>• Current research does not focus on family planning initiatives in smaller cities and urban peripheries<br><br>**Recommendations:**<br>• Future research on urban family planning needs to consider the neighbourhood effects, governance systems, migration and diversity of populations, and resilience strategies. |

| | | | | |
|---|---|---|---|---|
| Akwara et al. (2023) [12]<br><br>The Urban Environment and Disparities in Sexual and Reproductive Health Outcomes in the Global South: a Scoping Review | **SRH topics targeted in these interventions:**<br>SHRH programs and interventions adopted in LMIC urban settings<br><br>**Description of interventions in PHCs:**<br>**a)** Interactive text messages to improve retention of service users in HIV care; in addition to usual care (comprised of psychosocial support & counselling, education, screening and treatment) service users received weekly messages that they were asked to respond to within 48 hours.<br>**b)** Financial incentives in different modalities for HIV retesting - i) no incentives; ii) cash incentives for retesting at 3 and 6 months; iii) deposit contracts that would be lost if retesting is not done.<br>**c)** Cash transfers and combination HIV prevention interventions<br>**d)** Urban Reproductive Health Initiative (URHI) is a comprehensive family planning demand- and supply-side intervention, comprising of service quality, healthcare provider training, service user accessibility and affordability, and raising awareness of family planning methods. The program adopted context-specific approaches to address urban health inequities and ensure inclusive SRHR services for urban poor populations. | **Intervention a:** Autonomy/ privacy & confidentiality/ Communication<br>**Intervention b:** Autonomy/ supportive care<br>**Intervention c:** Autonomy/ supportive care<br>**Intervention d:** Dignity/ Autonomy/ Communication/ Health Facility Environment | • **Service user retention/follow up in care:** Intervention (a) did not have any impact on retention of HIV patients at the health facilities. Intervention (b) showed that service users who received the incentives were more likely to retest at 3 and 6 months compared to those who received no incentives or the deposit contracts.<br>• **Incidence of STIs:** Intervention (c) had no effect on incidence of herpes simplex virus type 2 at the 6, 12, and 18 month follow up visits (hazard ratio 0.96, 95% CI:0.67-1.38, P-value=0.83), however among rural low-risk women, those who received intervention (c), reported lower incidence of herpes (hazard ratio 0.45, 95%CI:0.29-0.71,P-value=0.001) compared to the control group.<br>• **Family planning uptake:** Intervention (d) increased uptake of family planning methods in urban cities in Kenya and Nigeria.<br><br>**Factors that may influence interventions:**<br>• Socio-cultural context<br>• The environment where urban individuals live in. | **Limitations:**<br>• Search was done on two databases and restricted to English language only.<br>• Inclusion of articles published after 2010<br>• No quality appraisal of studies was conducted due to heterogeneity of study designs.<br>• Review may not have identified all relevant studies due to the search terms used.<br><br>**Evidence gaps:**<br>• Evidence on SRHR interventions and policies in urban areas in the Global South is limited.<br>• Limited evidence on intra-urban disparities through comparisons between cities and through consideration of marginalized groups.<br>• There is a need to study how to effectively provide mHealth to reach urban poor and marginalized populations.<br><br>**Recommendations:**<br>• Further research focusing on how social issues e.g. poverty, inequality, social norms, intersect across different urban neighbourhoods and impact SRHR outcomes.<br>• Further research is needed to explore high |

| | | | | |
|---|---|---|---|---|
| | | | | risk and marginalized populations.<br>• Designing approaches and strategies tailored to the urban environment are necessary. |

| | | | | |
|---|---|---|---|---|
| Onaisi et al. (2022) [13]<br><br>Sexual risk behaviour reduction interventions in primary care in Organization of Economic Cooperation and Development countries. A systematic review | **SRH topics targeted in these interventions**: Counselling, behavioural and educational interventions related to reduction of risky sexual behaviours<br><br>**Description of interventions in PHCs:**<br>**a)** Very brief interventions (< 30minutes) that take place during a routine healthcare appointment. Described interventions were:<br>- educational intervention using socio-cognitive theories and theory of reasoned action<br>- Brief advice about safe sex and complementary resources<br>- 20 min intervention based on the social learning theory aimed at altering perception of risk for HIV, and increase knowledge and use of condoms.<br>- 10-20min intervention promoting condom use to service users with chlamydia<br>- Prevention counselling (RESPECT intervention) and rapid HIV testing in one session.<br>- brief patient-centred risk-reduction counselling and rapid HIV test<br>- Video (22 min long) added to usual care at STD clinic to promote condom use and reduce the number of partners using the Information, Motivation, Behaviour (IMB) model.<br><br>**b)** Brief interventions (30 - 60 minutes) that take place during prevention appointments<br>- Educational video played in the clinic waiting room regarding condom use addition to group discussions with role plays.<br>- Brief video played in the clinic waiting room showing STD prevention messages.<br>- HIV prevention intervention (VOICES/VOCES)<br>- One-on-one HIV/STD prevention counselling (RESPECT) with 2 sessions (40 minutes total)<br><br>**c)** Intensive interventions (>60 minutes and/or > 1 session)<br>- One-on-one HIV/STD prevention counselling with 4 sessions (200 minutes total)<br>- Intervention consisting of 1 of 4 versions of a | **Intervention a:** Autonomy/ communication/ supportive care<br>**Intervention b:** Autonomy/ communication/ supportive care<br>**Intervention c:** Autonomy/ communication/ supportive care<br>**Intervention d:** Autonomy/ communication/ supportive care | • **Condom use**: Very brief interventions (Intervention (a)) had mixed results but generally showed that these interventions did not have a significant impact on contraception use especially on the long run. Intervention (c - face-to-face approaches) showed no significant changes in contraceptive use; Intervention (c - group approaches) that were competency-based showing significant decreases in unprotected sexual intercourse. Intervention (d) showed that a combination of very brief and intensive approaches that were based on competencies resulted in more protected intercourses in intervention vs control, irrespective of the intervention duration.<br>• **Reduction in risky behaviours:** One of the approaches in intervention (c) (MAC-Choice Program) reported a significant reduction in the number of sexual partners compared to the control (1.53 vs 2.75;p-value=0.0001) and intercourse with occasional sexual partners compared to the control (0.26 vs 0.74;p-value=0.02) at 6 months follow-up.<br>• **Incidence of STIs:** Very brief interventions (Intervention (a)) did not have a significant effect on reduction in STI incidence. Brief interventions (Intervention (b)) showed a reduction in STI incidence generally; different studies of videos in waiting rooms reported reduction of incidence from 13.1% in control group to 6.3% in the intervention group (RR:0.49; p-value<0.05) and another study showed a small reduction in incidence of STIs compared to controls (4.9% vs 5.7%; HR0.9; 95%CI:0.84-0.99). The VOICES intervention (intervention b) showed lower incident STI rate after follow up (1.1% vs 13.5%, p-value<0.01). Face-to-face intensive interventions (intervention c) did not show significant results for the impact on STI. Group intensive interventions (Intervention c) had mixed results and were some showed a reduction in STI rates; PMI showed lower | **Limitations:**<br>• Most of the included RCTs in the review had high or concerning risk of bias.<br>• Grey literature search was not conducted.<br>• Meta-analysis was not conducted due to the heterogeneity of included studies.<br>• Findings need to be verified by conducting this research in more OECD countries.<br><br>**Evidence gaps:**<br>• Most interventions focused on high-risk adolescents and young adults, yet older adults (>50 years) are also sexually active but have lower use of contraceptives.<br><br>**Recommendations:**<br>• Interventions should consider a holistic approach to sexual health by not only focusing on risk reduction and disease prevention, but also the context and wellness of service users. |

| | | | |
|---|---|---|---|
| | 90-min risk-reduction individual counselling sessions that divided the IMB model using a full factorial approach.<br>- Behavioural intervention consisting of 5 weekly 60-90 minute individual sessions (MAC-Choice program) to reduce risky sexual behaviours.<br>- Four-session, individual, multi-component, cognitive/behavioural intervention aimed at preventing STDs.<br>- Group intervention for HIV/STD risk reduction consisting of one 250min session. The intervention sessions were either informative or competency-based.<br>- Four session group intervention using the IMB model to prevent STDs and HIV.<br>- An adapted evidence-based STI/HIV intervention (HORIZONS) and a Prevention, Maintenance Intervention (PMI) consisting of 4 hour group sessions followed by brief telephone contacts every 8 weeks for 36 months to complement prevention messages.<br>-HIV/AIDS risk reduction intervention with five group sessions focusing on risk education; skills training in condom use, sexual assertiveness, problem solving, and risk trigger self-management; and peer support for change efforts.<br>- Behavioural intervention for small groups consisting of seven sessions (90-120min each) about HIV risk reduction.<br>- Behavioural intervention with three small-group sessions of 3-4 hours each aimed at helping women recognize personal susceptibility, commit to behavioural change, and acquire skills.<br><br>**d)** Combination of brief and intensive interventions<br>- a combination of one of two brief interventions (individually delivered and based on either informative or motivations interviewing) and one of two intensive interventions (group delivered and based on informative or IMB models)<br>- combination of very brief and intensive sessions | | cumulative incidence of chlamydia over 36 months (aRR=0.5,p-value=0.02). Intervention (d) showed that a combination of very brief and intensive approaches that were based on competencies resulted in lower STI prevalence after 12 months, irrespective of the intervention duration. |

| | |
|---|---|
| | based on two approaches, informative or competency-based. |

| | | | | |
|---|---|---|---|---|
| Mariño et al. (2023) [14]<br><br>Educational interventions for cervical cancer prevention: a scoping review | **SRH topics targeted in these interventions:** Educational interventions for cervical cancer screening<br><br>**Description of interventions in PHCs:**<br>**a)** Group discussion sessions where women asked questions about cervical cancer prevention and voiced their concerns.<br>**b)** Teaching interventions that were based on lectures, 'expository lessons' or 'card games'.<br>**c)** Interventions of resource support, complementary to other interventions, using booklets, leaflets, and informative videos with educational content.<br>**d)** Telephone interviews to provide education on cervical cancer and possible risks, importance of pap smear tests, continuity of testing, care and follow-ups.<br>**e)** Peer-Led Navigation (PLNav) intervention aimed to provide health service access by selecting providers from the same communities as the women and speak the same language or from the same context. | **Intervention a:** Dignity/ communication, supportive care<br>**Intervention b:** Autonomy/ communication<br>**Intervention c:** Autonomy/ communication/ confidentiality & privacy<br>**Intervention d:** Autonomy/ communication/ supportive care<br>**Intervention e:** Dignity/ communication, supportive care | • **Cervical cancer and screening knowledge:** Intervention (c), especially videos, reported increase in cervical cancer knowledge. Intervention (e) showed that participating women had increased knowledge on cervical cancer screening.<br>• **Pap smear uptake:** Intervention (c), specifically videos, was found to increase confidence in women to schedule pap smears. Intervention (d) led to increased attendance at pap smear health services. Overall, the review found that educational interventions that were given by community health workers were very well received by participating women.<br><br>**Factors that may influence interventions:**<br>• Theory-based and culturally tailored interventions and approaches. | **Limitation:**<br>• References of selected studies were not reviewed for other eligible studies.<br><br>**Evidence gaps:**<br>• Described educational programs had a low participation of nurses<br><br>**Recommendation:**<br>• Future research needed to assess whether including nurses will lead to alleviating barriers in health prevention behaviours. |
| Atere-Roberts et al. (2020) [15]<br><br>Interventions to increase breast and cervical cancer screening uptake among rural women: a scoping review | **SRH topics targeted in these interventions**: breast and cervical cancer screening<br>**Description of interventions in PHCs:**<br>**a)** Friend-to-friend and Patient Navigation (FTF+PN) program consists of 'Pink Parties' by cancer prevention specialists to educate underserved, uninsured, older women about breast an cervical cancer screening. The 'parties' are made up of 1- oral presentation of mammography benefits by clinical staff, 2- facilitator-led small group discussion about mammography and concerns, 3- a service user-facilitator session to help the service user ask their providers for mammography screening.<br>**b)** Ohana day project is a one-day program consisting of one-on-one visit with a physician for screening and education about cancer, as well as provision of culturally appropriate brochures | **Intervention a:** Autonomy/ dignity/ communication/ supportive care<br>**Intervention b:** Autonomy/ dignity/ communication/ supportive care | • **Mammography or pap smear uptake:** Intervention (a) had mixed results depending on race/ethnicity; English-speaking Latina women had lower odds of receiving a mammogram (OR:0.60; 95%CI:0.43-0.83) and Pap test (OR:0.66; 95%CI:0.47-0.92) than non-Hispanic White controls; Spanish-speaking Latinas had higher odds of Pap test uptake (OR:1.64; 95%CI:1.22-2.20) compared to same controls. Intervention (b) reported improvements in breast cancer screening, where 84% of women who received the intervention had a mammography at the 6-month follow-up compared to 66% of the comparison group (p-value=0.002).<br><br>**Factors that may influence interventions:**<br>• Tailoring interventions to specific groups of people or contexts<br>• Administrative costs | **Limitations:**<br>• The review search strategy was restricted to PubMed/Medline database.<br>• Potential publication bias<br>• Only peer-reviewed studies were included<br>• Strict inclusion criteria, especially in relation to 'rurality'.<br>• Comparison for interventions between studies was not possible due to high heterogeneity<br><br>**Evidence gaps:**<br>• Minimal research evaluating interventions' association with |

| | | | | |
|---|---|---|---|---|
| | | | • Assistance in scheduling appointments<br>• Travel distance, transportation difficulties, and access to specialty care. | increased community access to cervical cancer screening.<br>• No studies described interventions that emphasized repeated screening.<br>• There was limited geographic diversity and representation of underserved/minority populations, within USA, in the eligible studies.<br><br>**Recommendations:**<br>• Future research should look at interventions for on-time repeat screening procedures, especially in rural areas.<br>• Researchers need to define more explicitly what 'rural' indicates. |
| Aung et al. (2017) [16]<br><br>Interventions for Increasing HIV Testing Uptake in Migrants: A Systematic Review of Evidence | **SRH topics targeted in these interventions**: HIV testing uptake among migrants<br><br>**Description of interventions in PHCs:**<br>**a)** Women's Health Program (WHP) is an interactive HIV education program that consists of general women's health education about HIV transmission and prevention, sexual and reproductive autonomy, condom practice and negotiation skills.<br>**b)** Provider-initiated HIV testing and counselling (PITC) involved general practitioners proactively proposing HIV testing and counselling to migrants.<br>**c)** HIV-related educational announcements and invitation for free HIV testing for service users while they waited in the waiting area of the outpatient departments. | **Intervention a:** Autonomy/ communication/ supportive care<br>**Intervention b:** Communication/ supportive care<br>**Intervention c:** Dignity/ autonomy | • **HIV testing Uptake:** Intervention (a) compared to the control group had significantly higher odds of HIV testing after 12 weeks (OR: 2.50; 33% change) but no significant change was recorded 3 months post intervention. Intervention (c) led to an increase in HIV testing among the clinic clients from 6.7% during standard care to 25.4% during intervention period; and the mean number of HIV-positive clients per week rose from 0.9 during standard care to 5.6 during the intervention period.<br>• **Feasibility and acceptability of intervention**: Intervention (b) was accepted by service users, yet physicians stated discomfort when asking for HIV testing during an irrelevant appointment and routine implementation was not feasible all the time due to time pressure and length of the counselling intervention. | **Limitations:**<br>• Only peer-reviewed studies were included.<br>• Some HIV testing interventions may have been missed.<br>• The search strategy focused on international migrants, yet ethnic minorities may include migrants and would have been missed.<br>• Due to the presence of qualitative studies, the risk of bias tool had to be modified to accommodate this.<br><br>**Evidence gaps:** |

| | | | | |
|---|---|---|---|---|
| | | | **Factors that may influence interventions:**<br>• Resource intensive interventions that affect scalability.<br>• Stigmatization and discrimination of vulnerable groups.<br>•Provider concerns about implementation | • Minimal research on HIV testing among migrants outside of USA, while most migrants are in Asia and Europe.<br><br>**Recommendations:**<br>• A more thorough search of the grey literature and contact of organizations is necessary.<br>•Further research is needed for different migrant groups and settings, and considering not only the health seeking behaviours but also the migration status, health system determinants and the economic aspects of HIV testing. |
| Sorhaindo et al. (2022) [17]<br><br>Interventions to reduce stigma related to contraception and abortion: a scoping review | **SRH topics targeted in the interventions:** 1 on contraception and abortion education and 1 on abortion experiences.<br><br>**Description of interventions in PHCs:**<br>**a)** Family Planning Balanced Counselling Strategy (FP-BCS) to support uptake of modern contraception, through practical, interactive counselling that supports clients to make their decisions.<br>**b)** Narrative intervention using a 4-minute short film screened at clinic waiting room, before receiving care, to normalize the abortion experience. | **Intervention a:** Dignity/ autonomy/ privacy & confidentiality/ communication/ Supportive care<br>**Intervention b:** Dignity/ communication/ Supportive care/ health facility environment | • **Contraceptive knowledge:** The FP-BCS intervention (a) improved attitudes toward modern contraception.<br>• **Abortion internalized stigma:** Intervention (b) showed no significant difference in internalized stigma between intervention and control groups. | **Limitations:**<br>• Search limited to studies in English only.<br>• Studies that did not evaluate intervention and only described activities were excluded.<br>• Publication bias favouring studies showing effectiveness only.<br><br>**Evidence gaps:**<br>• There is a limited number of interventions tackling reproductive health stigmas.<br>• Most of the available literature originates from the USA with little knowledge about other contexts. |

| | | | | |
|---|---|---|---|---|
| | | | | • A limited number of interventions deal with stigma reduction at the structural level, as opposed to the individual level.<br><br>**Recommendations:**<br>• Future research should develop and test theory-based interventions and use validated scales to measure their effectiveness.<br>• Based on the results of this scoping review, future systematic reviews can focus on effectiveness of interventions. |
| AlHamawi et al. (2023) [18]<br><br>Family planning interventions in Jordan: A scoping review | **SRH topics targeted in the interventions:** effective use of oral contraceptive pills.<br><br>**Description of interventions in PHCs:**<br>**a)** An information-based booklet on effective use of oral contraceptive pills was provided to women who used the pill and attended pharmacies or community/fertility /OBGYN clinics. The booklet contained information on types or oral contraceptives, mechanisms of action, precautions, advantages/disadvantages of each type, optimal use guidance and information about alternative contraceptive methods. | **Intervention a:** Dignity/ autonomy/ communication/ Supportive care | **• Contraceptive - oral pills - knowledge:** Intervention (a) significantly improved knowledge (from 1.76 to 5.00 post-intervention) and maintained a high score at follow-up (4.93), compared to no significant change in control group women. Attitude score also significantly improved among women in the intervention group (from a score of 5.10 to 5.50 post intervention), compared to no change in the control group.<br><br>**Factors that may influence interventions:**<br>• misconceptions and social norms about family planning. | **Limitations:**<br>• Only one database was used for the search.<br>• The search strategy may have not been comprehensive enough.<br><br>**Evidence gaps:**<br>• Limited evidence on High Impact Practices (HIP), which are evidence-based practices; and on enhancements of HIPs such as family planning vouchers, adolescent targeted services or digital healthcare.<br><br>**Recommendations:**<br>• Family planning interventions need to be designed with positive |

| | | | | |
|---|---|---|---|---|
| | | | | messages to encourage contraception use and address misconceptions and biases.<br>• Interventions should target parents with family planning information, which can be transferred to young people.<br>• Digital mHealth interventions should be explored. |

| | | | | |
|---|---|---|---|---|
| Boydell et al. (2022) [19]<br><br>Getting Intentional about Intention to Use: A Scoping Review of Person-Centered Measures of Demand | **SRH topics targeted in these interventions:**<br>Interventions related to intention to use contraception and family planning<br><br>**Description of interventions in PHCs:**<br>**a)** A 15 minute 'Health Talk' by about family planning, given by community clinic health assistants.<br>**b)** A contraceptive behavioural intervention delivered by mobile phones text messaging. The messages were tailored according to marital status and the content included information about contraception, its misconceptions and side effects.<br>**c)** Counselling messages for HIV risk and use of injectable contraceptive methods | **Intervention a:**<br>Autonomy/ communication/ supportive care<br>**Intervention b:**<br>Autonomy/ dignity/ privacy & confidentiality<br>**Intervention c:**<br>Autonomy/ communication/ supportive care | • **Intention to initiate a new family planning method**: Intervention (a) led to 33% of participants (45% of women participants) reporting that they wanted to try a new method. Intervention (b) increased agreement on intent to use effective contraceptive methods. Intervention (c) did not lead to a change in use of injectable contraceptives, yet 60% of participants stated they would use a condom for dual protection.<br>• **Contraception knowledge:** Intervention (b) led to a moderate improvement of effective contraception knowledge among Palestinian women compared to the control group.<br><br>**Factors that may influence interventions:**<br>• Socioeconomic status<br>• Behavioural variables such as the individual's knowledge and attitudes. | **Limitations:**<br>• Two databases were used in the search only.<br>• The search was limited from 1990 to 2020.<br><br>**Evidence gaps:**<br>• Research about the definition and measurement of 'Intention to Use' (ITU) construct is limited.<br><br>**Recommendations:**<br>• Further work is needed to develop a comprehensive measure of ITU, explore how this measure varies in relation to long-term goals vs short term ones, and examine how ITU affects contraception use. |

| | | | | |
|---|---|---|---|---|
| Footman et al. (2021) [20]<br><br>A Systematic Review of New Approaches to Sexually Transmitted Infection Screening Framed in the Capability, Opportunity, Motivation, and Behavior Model of Implementation Science | **SRH topics targeted in these interventions**: Sexually transmitted infections screening approaches<br>**Description of interventions in PHCs:**<br>**Electronic medical records interventions:**<br>**a)** Changes in testing policy on the computer laboratory sheet order from 'opt in' to 'opt out'<br>**b)** Electronic health record decision support tool which provides alerts for sexually active females presenting to the clinics who did not have annual screening.<br>**Sample Collection interventions:**<br>**c)** Self-testing program without need for appointment or offered by clinician at the end of consultation<br>**d)** Express testing criterion intervention for low-risk individuals to skip physical examination and receive STI testing<br>**e)** Test-and-Go (TAG) service where nurses collect blood and throat swabs, service user collects urine and rectal samples privately and drops off at clinic<br>**Improved Technologies**<br>**f)** Better Point-of-care (POC) testing using new diagnostics - NAAT POC same day testing and diagnosis<br>**g)** NAATs POC technologies with 30-min 90-min and 2hour waiting times for diagnosis results<br>**h)** NAATs POC tests with sample first approach - service user testing is done before their consultation.<br>**Treatment accuracy - Antimicrobial Stewardship**<br>**i)** AMR marker testing | **Intervention a:** supportive care<br>**Intervention b:** Communication/ supportive care<br>**Intervention c:** Autonomy/ dignity/ confidentiality & privacy<br>**Intervention d:** Supportive care<br>**Intervention e:** Dignity/ supportive care<br>**Intervention f-g-h:** Supportive care<br>**Intervention i**: Supportive care | • **STI screening uptake:** Intervention (a) increased STI testing rate from 5.5% to 45.2%. Intervention (b) resulted in adolescent girls being 2.1 times more likely to be screened for chlamydia. Intervention (c), in one study, showed an increase of 32% in pharyngeal testing (444 to 586 tests) and 33% increase in rectal testing (390 to 520 tests); additional results showed a increased yield of detection of chlamydia (47%) and gonorrhoea (50%) infections when compared with baseline data.<br>• **Diagnosis and treatment:** Intervention (d), compared to standard care, led to a significant rise in the diagnosis of chlamydia, gonorrhoea, and syphilis infections. Intervention (e) had no impact on providing earlier treatment. Intervention (f) led to lower re-infections from 20% before the intervention to 12% after the intervention (prevalence ratio, 0.60; 95% CI, 0.33–1.09) and reduced the number of times service users needed to visit the clinic to get tested and then treated. Intervention (g) led to increase in appropriate treatment from 52% to 100% compared to lab-based testing; and another study stated that intervention (g) corrected treatment decision 20% of the time, avoiding unnecessary treatment.<br>• **Preference and attitudes:** Service users reported that intervention (c)'s adoption increased their testing frequency and that the self-instructional pictures for guidance were very helpful. In another intervention (c) implementation, providers stated it saved time in paperwork and sample collection and led to service user privacy and uncreased testing; yet service users found the instructions unclear and led to confusion of the process. Intervention (g) waiting time acceptance varied by different service users and depended on circumstances, hence testing acceptability first is important before implementation. Intervention (h) was | **Limitations:**<br>*None identified*.<br><br>**Evidence gaps:**<br>*None identified*.<br><br>**Recommendations:**<br>• When developing new interventions, it is important to consider the service users.<br>• Further evaluations of STI screening services are necessary to identify what works and what needs to be revised. |

| | | | |
|---|---|---|---|
| | | | accepted by 90% of participants who were surveyed, stated that they were willing to wait 1-2 hours for results, however, more than 2 hours was not acceptable. Intervention (i) waiting time was not always accepted by service users unless they were concerned about specific infections.<br><br>**Factors that may influence interventions:**<br>• Clear instructions and guidelines are necessary to ensure proper use of the self-service<br>• Availability of the instruments and space at the PHCs.<br>• Staff availability.<br>• STI education and knowledge among service users |

| | | | | |
|---|---|---|---|---|
| Munyuzangabo et al. (2019) [21]<br><br>Delivery of sexual and reproductive health interventions in conflict settings: a systematic review | **SRH topics targeted in these interventions:**<br>Main components of SRH: HIV and STIs, gender based violence, family planning, and general SRH interventions offered to refugees and internally displaced populations within camps and outside camps.<br><br>**Description of interventions in PHCs:**<br>**Family planning services:**<br>**a)** Contraception provision<br>**b)** Abortion and post-abortion care<br>**c)** Counselling<br>**d)** Behavioural education<br>**e)** Screening interventions for referral<br>**HIV/STI interventions:**<br>**f)** HIV/STI prevention, treatment and follow up<br>**g)** HIV/STI screening for referral<br>**h)** Behavioural education<br>**i)** Counselling<br>**Gender-based violence interventions:**<br>**j)** Counselling<br>**k)** Behavioural education<br>**l)** Emergency contraception | **Intervention a:** Supportive care<br>**Intervention b:** Autonomy/ dignity/ communication/ supportive care.<br>**Intervention c:** Autonomy/ communication/ supportive care<br>**Intervention d:** Autonomy/ communication/ supportive care<br>**Intervention e:** communication/ supportive care<br>**Intervention f:** Autonomy/ dignity/ communication/ supportive care<br>**Intervention g:** Communication/ supportive care<br>**Intervention h:** Autonomy/ communication/ supportive care<br>**Intervention i:** Autonomy/ communication/ supportive care<br>**Intervention j:** Autonomy/ communication/ supportive care<br>**Intervention k:** Autonomy/ communication/ supportive care<br>**Intervention l:** | Only 18% of studies included in this review reported on intervention coverage and 7% reported on intervention effectiveness.<br><br>• **Contraceptive prevalence rate:** Most studies reported rates ranging from 1% to 39.6%. There was a variation in uptake by country, for example, LARC varied from 1% in Djibouti to 78% in DR Congo. Compared to Intrauterine devices and other modern methods, implants were most commonly accepted in Chad and DR Congo.<br>• **Provision of Emergency Contraception:** Uptake ranged from 50% (NGO services) to 90% (healthcare professionals).<br>• **Family planning counselling uptake:** Counselling post-abortion coverage ranged from 35%-98%.<br>• **Effectiveness of counselling and testing services**: When offered vouchers for free services, there was a higher uptake among IDP women compared to non-IDP women in surrounding communities.<br><br>**Factors that may influence interventions:**<br>• Barriers: security and logistical issues, lack of funding and resources, lack of training and presence of skilled health workers, women's inability to access healthcare due to limited mobility or financial constraints, social norms and stigma.<br>• Enablers: collaboration between NGOs, community and Ministry of Health, presence of community health workers, community engagement, culturally appropriate interventions | **Limitations:**<br>• The grey literature search was limited to a selection of NGO websites and may not have captured all interventions.<br>• The search was restricted to English only.<br>• published literature may contain limited details about interventions in conflict-affected settings.<br><br>**Evidence gaps:**<br>• Limited literature on displaced populations residing outside the camps.<br>• Scarce research on SRH interventions targeted towards adolescents.<br>• MISP guidelines for interventions were not followed explicitly.<br>• Interventions related to safe abortion or post-abortion care were limited.<br>• Other SRH issues e.g. cancers, obstetric fistula, were barely mentioned.<br>• No research on intervention effectiveness related to SRH outcomes.<br>• Gender-based violence interventions were described in grey literature mainly rather than peer reviewed articles.<br><br>**Recommendations:** |

| | | | | |
|---|---|---|---|---|
| | | Dignity/ supportive care | | • Further research required for identifying how to engage community-based individuals to deliver selected SRH interventions.<br>• For more effective interventions, involvement of community leaders is crucial.<br>• The modality of intervention delivery, whether in clinics or outreach needs to be assessed.<br>• Further research is necessary for displaced populations outside camps and for adolescents.<br>• More research is required to assess SRH intervention effectiveness in conflict settings.<br>• Qualitative research can help identify guidelines that humanitarian actors use to develop interventions. |

SRH: Sexual and Reproductive Health; SRHR: Sexual and Reproductive Health Rights; PHC: Primary Care Setting; IUD: Intrauterine device; FP: family planning; RH: Reproductive health; PCC: person-centred care; STI: Sexually transmitted infection; LARC: Long-Acting Reversible Contraceptive; OR: odds ratio; RR: relative risk; aRR: adjusted relative risk; HR: Hazard rate; CI: Confidence interval; LMIC: Low- and Middle-Income Country; IDP: Internally-displaced population; NGO: Non-governmental organisation
